# Global trends and prevalence of scabies: a spatiotemporal analysis using Global Burden of Disease 2021 data

**DOI:** 10.1101/2025.01.24.25321000

**Authors:** Saptorshi Gupta, Simon Thornley, Arthur Morris, Gerhard Sundborn, Cameron Grant

## Abstract

**Background:** Scabies is a neglected disease believed to be more prevalent in resource-poor nations. Data describing global temporal trends of scabies incidence and prevalence and risk factors is limited.

**Methods:** This paper uses joinpoint regression and spatial analysis to estimate age-standardized rates (ASR) of scabies incidence and prevalence and temporal trends in 204 countries and regions from 1990 to 2021. Local indicators of spatial association were used to determine contiguous areas of high prevalence. The association of socio-demographic and economic factors with scabies was determined using locally weighted scatterplot smoothing and log-normal regression models.

**Findings:** Global prevalence of scabies in 2021 was 2.71% (95% confidence interval [CI]: 2.41 to 3.04%). Age-standardized rates of scabies have marginally declined globally from 1990 to 2021 with an Average Annual Percentage Change (AAPC) of -0.10 (95% CI -0.05 to -0.14) for incidence and -0.09 (95% CI -0.05 to -0.14) for prevalence. Spatial clustering of high scabies prevalence was present in tropical Latin America, Southeast Asia, and the Pacific Islands. Rates have shown an increasing trend in high-income regions such as Australasia and parts of Europe. Scabies showed a declining trend from middle to high socio-demographic index (SDI) regions. There is a significant positive association between warmer latitudes and increasing urbanization and scabies prevalence.

**Interpretation:** Scabies prevalence remains high in many regions. Control programmes should be prioritized to accelerate progress to reduce prevalence of this important neglected disease.

## Background

Caused by *Sarcoptes scabiei*, scabies is an infectious disease that presents with symptoms of itch and rash, with secondary bacterial infections (1) or impetigo frequent complications (2). The need to relieve itch often leads to excoriation (1) which can become infected with *Streptococcus pyogenes*, in-turn resulting in further complications including post-streptococcal glomerulonephritis (3), acute rheumatic fever (ARF) and rheumatic heart disease (RHD) (4,5). Since the health and economic burden of scabies is substantial, it is important to understand its distribution and determinants at the global, national and regional levels.

The World Health Organization (WHO) estimates the total global count of scabies cases to be at 200 million at any given time (6). Its prevalence is disproportionately higher in tropical regions, especially among children, elderly and the immunocompromised (1). Despite its considerable health impacts and high prevalence, scabies remains a relatively neglected disease with mass-drug administration (MDA) programs implemented often sub-nationally (7), rather than being co-ordinated globally.

Understanding the distribution and determinants of scabies and its changing temporal trends could help guide disease control policy. Estimates of spatiotemporal trends of scabies at the global, national and regional level and risk factors for scabies are limited. This study uses data from the Global Burden of Diseases (GBD) to derive these estimates from 1990 to 2021 and identify the high-risk areas where prevalence is increasing.

### Data and Methods

#### GBD Study

The GBD study is a detailed and comprehensive data collection that estimates 459 risk factors and health conditions both globally and nationally (8). Estimates of incidence, prevalence, mortality, years of life lost (YLLs), years of life lived with disability (YLDs) and disability-adjusted life years (DALYs) are reported for each condition.

For this study, the incidence, prevalence and age-standardized rates of incidence (ASIR) and prevalence (ASPR) for scabies was obtained by sex and age group for 21 regions, 204 countries or territories for 1990 to 2021 from the online GHDx data source query tool (http://www.healthdata.org/gbd/), where the details of GBD data collection are described. In case of missing values, data from neighbouring areas were used as estimates to impute data (9).

Countries were categorized based on region, income and sociodemographic index (SDI) – a composite index of development based on lagged per capita income, average level of education for populations aged 15 years and above and, total fertility rates for women less than 25 years old. Sociodemographic index values range from 0 (poorest) to 1 (wealthiest) with this index used to evaluate the development status of each location year. Countries were classified into five development quintiles based on the 2021 GBD data: low, low-middle, middle, high-middle and high SDI regions (8).

### Scabies definition

The definition of scabies was based on the International Classification of Diseases (ICD) diagnostic criteria. Scabies cases were identified from ICD-10 coded B86 cases from outpatient clinic data and a comprehensive literature review which included surveys or other published estimates of the frequency of disease.

### Data Sources

For the GBD data collection, scabies data was extracted from a comprehensive literature review of previously published studies, USA claims data, outpatient data and USA MarketScan 2000 data. There has been a noteworthy change in modelling and data sources available compared to the previous GBD estimates. USA claims data from 2000 and 2010 through 2016 and outpatient data were included and for modelling, within-DisMod crosswalks were replaced with crosswalks completed using the MR-BRT modelling tool (10).

Five covariates – urban population as a percentage of total population, population density (number of people per square kilometre of land area), life expectancy at birth (LEB), unemployment as a percentage of total labour force, and Gross National Income (GNI) per capita by purchasing power parity (PPP) 2021 were collected from the DataBank of the World Bank’s World Development Indicators (11).

### Statistical analyses

To facilitate comparison among several countries, age-standardized rates (ASR) of scabies incidence and prevalence were directly collected from the GBD database and analyzed using R (12) and QGis (13). In the GBD analysis, incidence refers to the number of new count cases during a specific time period in a particular population, the rates measured by the number of new cases in a year divided by the mid-year population. Prevalence, on the other hand, refers to the total number of cases in a particular population over a period and prevalence rate refers to the proportion of the number of cases divided by the mid-year population. All estimates in this GBD study refer to point prevalence. Temporal changes in rates were calculated using Joinpoint regression software (14). Trends were determined by calculating the Average Percentage Change (APC) and Annual Average Percentage Change (AAPC), a summary widely used to measure ASR trends. Average Percentage Change was calculated by taking the geometric mean of scabies rates for each year and fitting a straight line with this geometric mean as the dependent variable and year as the independent variable.

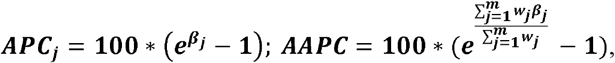

Where *β*_*j*_ : slope of *j*th time segments; : *w*_*j*_ is the length of the *j* th time segment. A minimum of 0 and maximum of 6 join points were selected using the Grid Search method over the study period. The permutation test was used for modelling. A parametric method was used to calculate the confidence intervals for APC and AAPC. A two-sided *P*-value of less than 0.05 indicated that APC and AAPC were significantly different from the null value (zero), representing no change. Trends are considered increasing when both AAPC and ASR are greater than zero and decreasing when both are less than zero. The association of temporal trends in prevalence rates in major regions and socio-demographic index (SDI) was determined using Locally Weighted Scatterplot Smoothing (LOWESS). Global Moran’s *I* and Local Indicators of Spatial Association (LISA) or Local Moran’s *I* was used to detect clusters of high scabies prevalence between neighbouring countries. Moran’s *I* ranges from -1 (perfect dispersion) to 1 (perfect clustering). A log-linear regression model was used to determine the association between scabies rates and socioeconomic and geographic factors. Absolute values of latitude were taken to consider the equator as a baseline and latitude on either side as equal. Right skewed data was log-transformed to restore normality and multicollinearity between covariates were assessed before performing the regression analysis. A variance inflation factor (VIF) of more than 10 was considered substantial evidence of multicollinearity.

A conceptual framework of the data collection process and statistical analysis has been enumerated in Supplementary Figure 1.

## Results

### Global distribution of scabies burden

Globally, the total cases of scabies incidence increased by 34.6% for both sexes from 462 million cases (95% CI: 407 million to 526 million) in 1990 to 622 million (95% CI: 556 million to 694 million) in 2021 (**Table 1**). However, the age-standardized incidence rate (ASIR) of scabies showed a significant decrease of 2.7% over this period from 8.27 per 100 (95% CI: 7.32 to 9.31) in 1990 to 8.05 per 100 (95% CI: 7.17 to 9.02) in 2021, with an average annual percentage change (AAPC) of -0.10 (95% CI: -0.05 to -0.14).

**Table 1:**
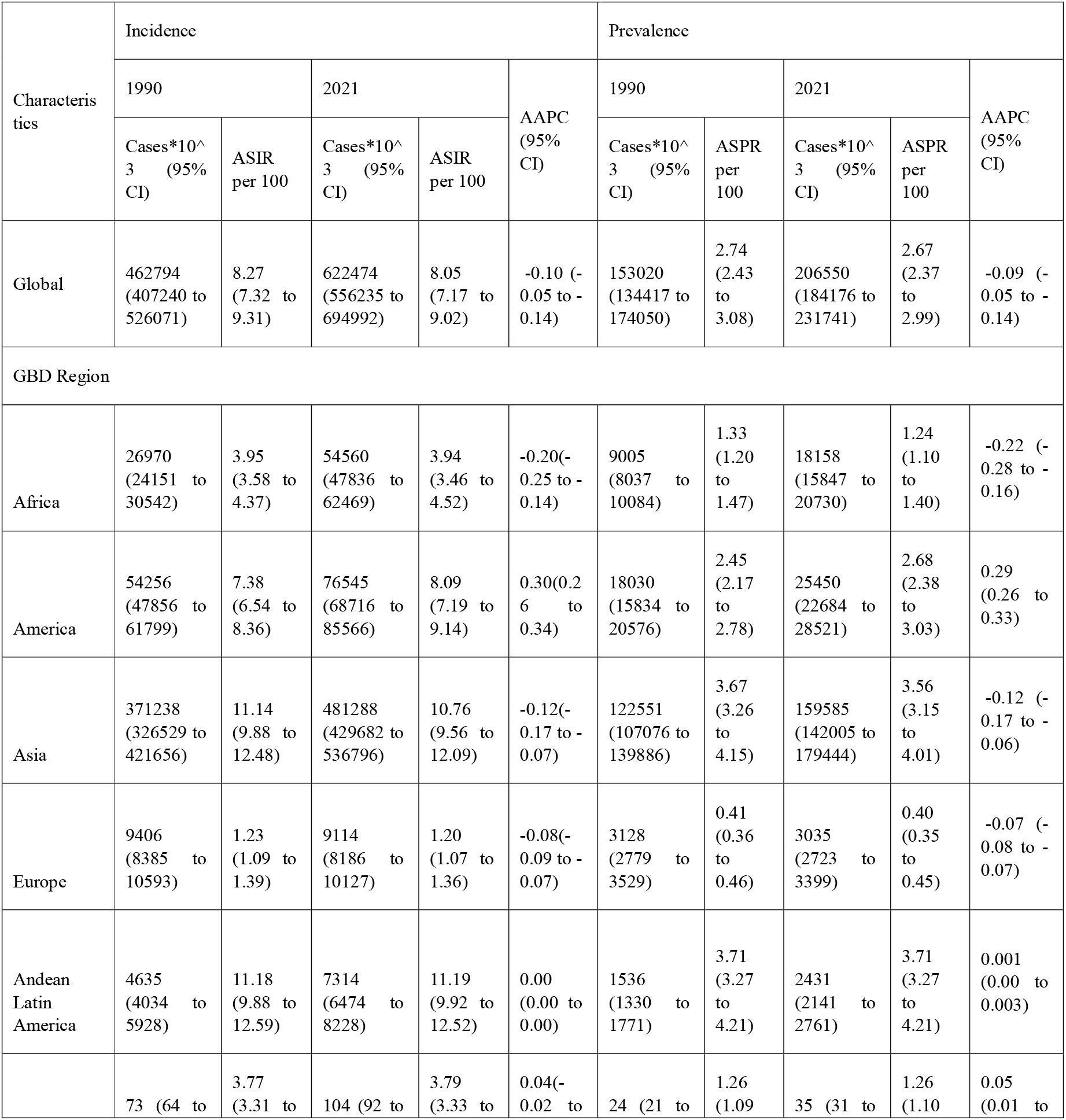
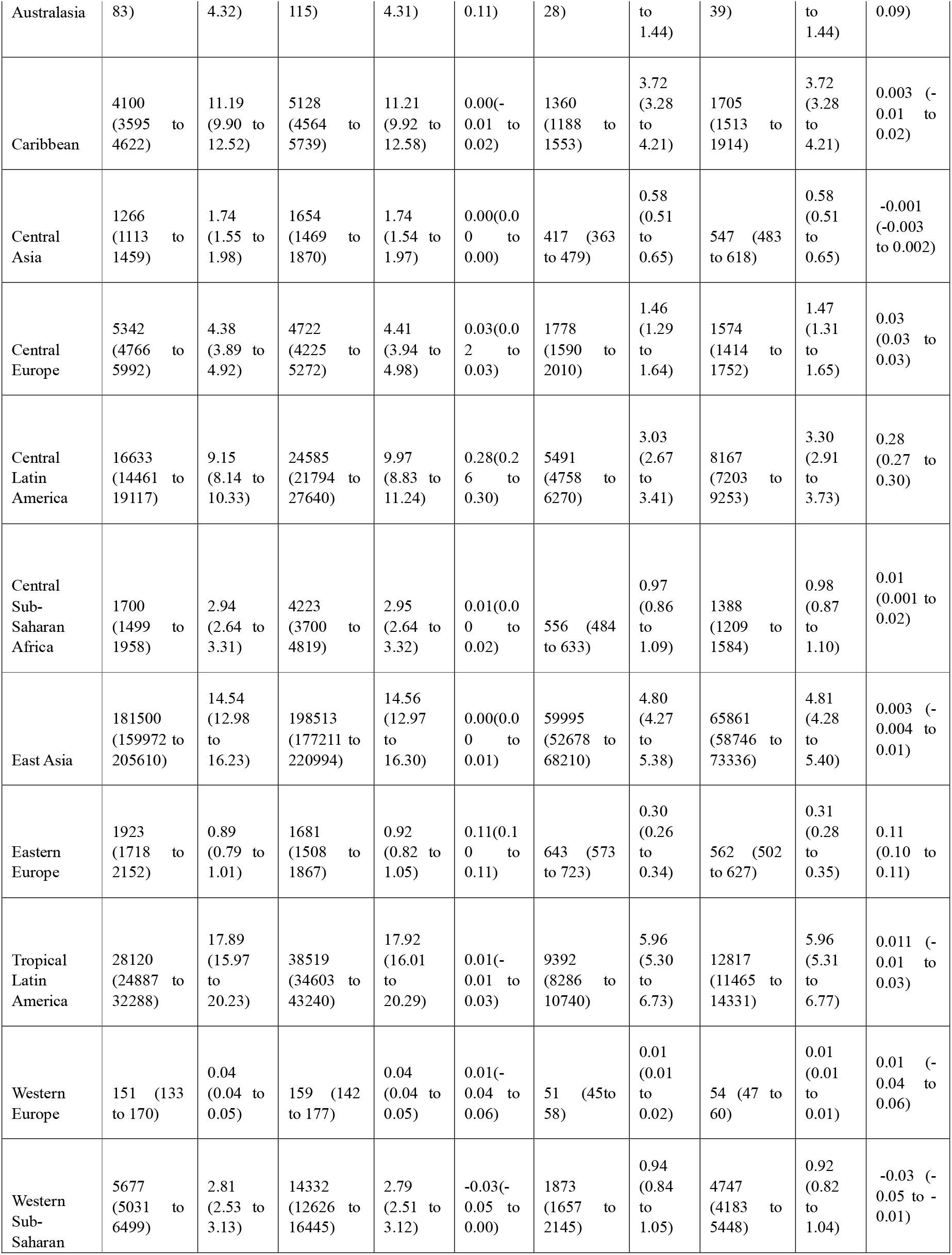
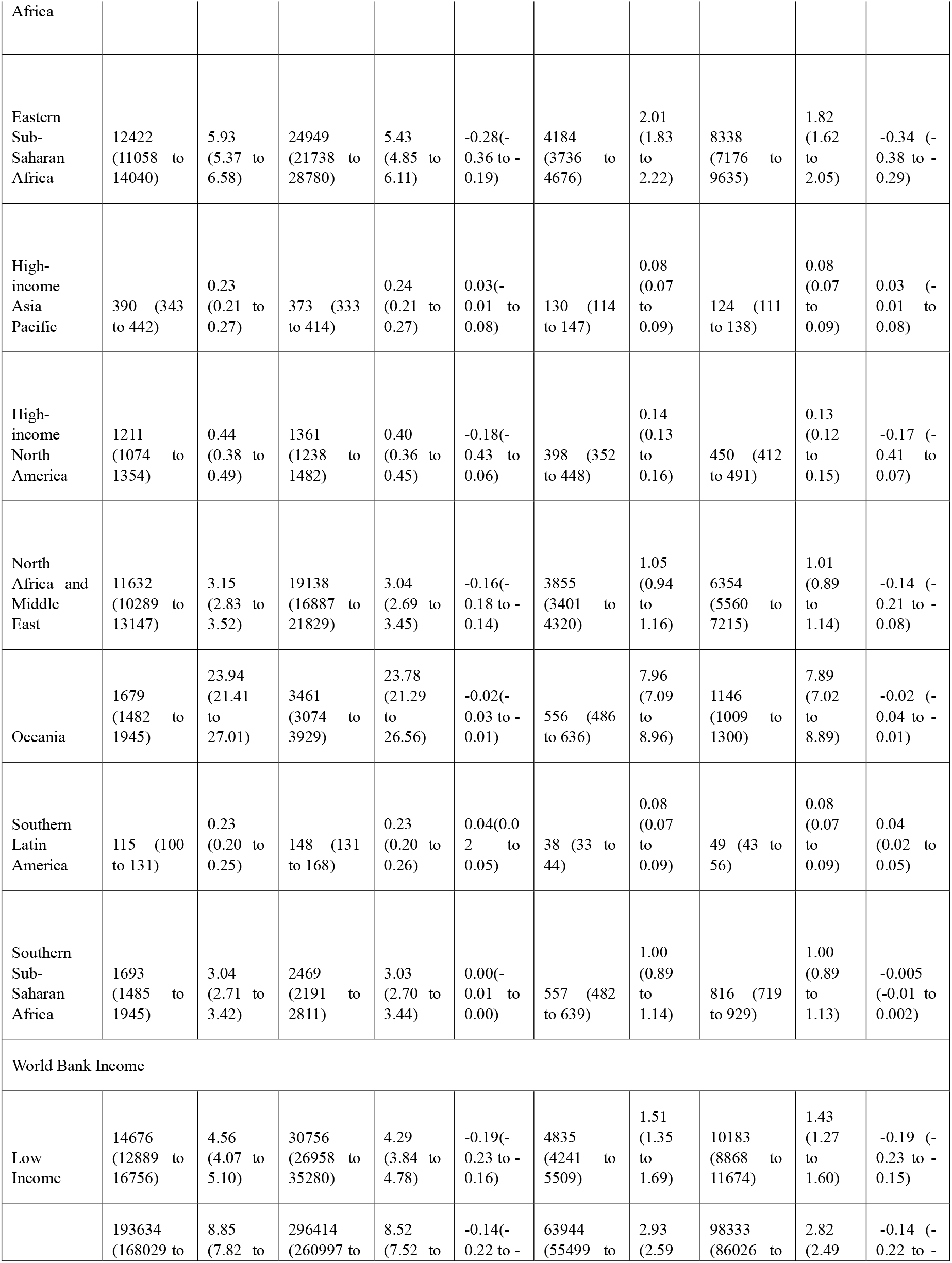
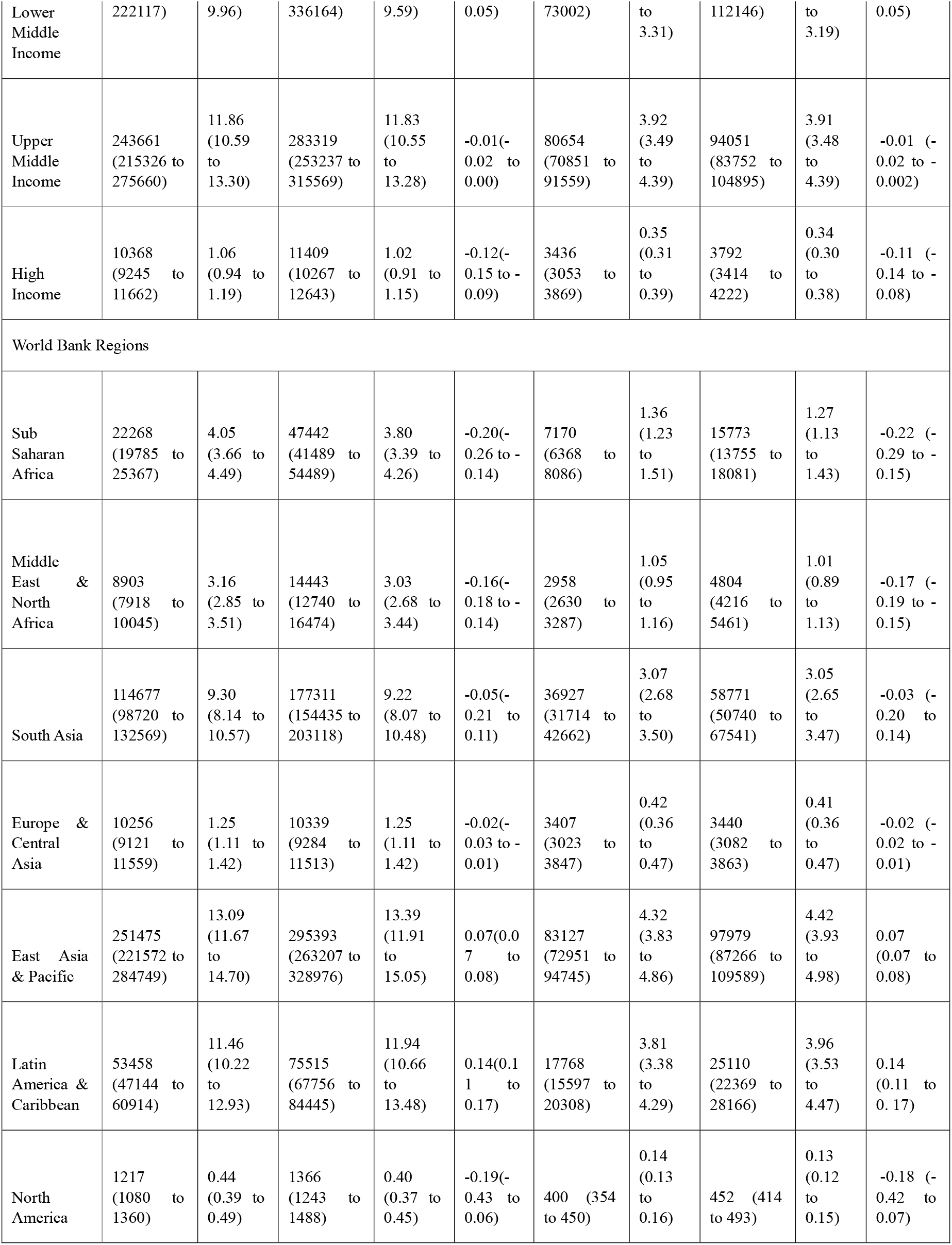
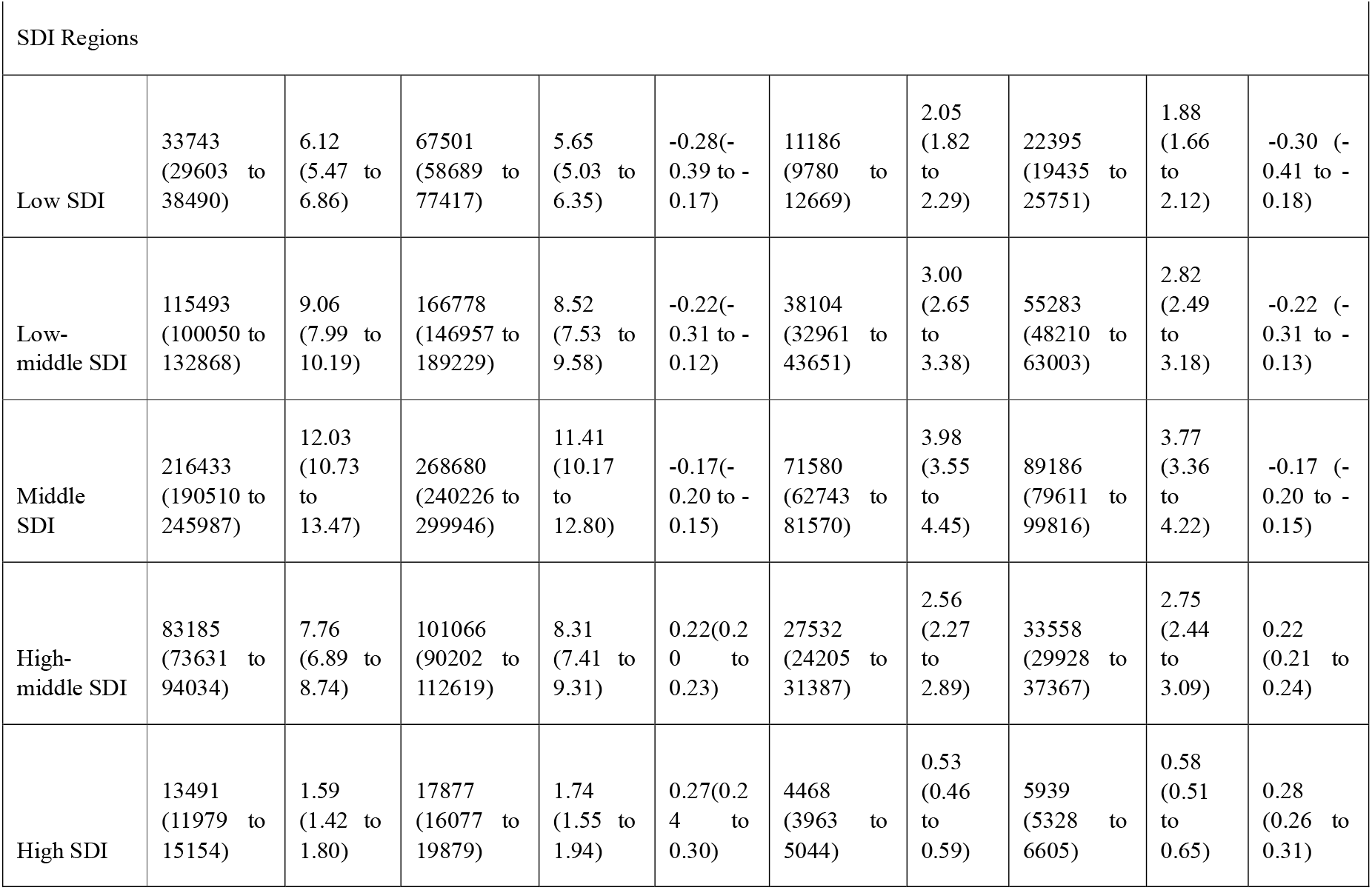
Incidence and prevalence of scabies and its rate of change from 1990 to 2019 in major parts of the world.

Scabies accounted for about 153 million cases (95% CI: 134 million to 174 million) in 1990 and 206 million cases (95% CI: 184 million to 231 million) in 2021. The age-standardized prevalence rate (ASPR) decreased significantly from 2.74 per 100 (95% CI: 2.43 to 3.08) in 1990 to 2.67 per 100 (95% CI: 2.37 to 2.99) in 2021, with an AAPC of -0.09 (95% CI: -0.05 to -0.14).

The ASIR and ASPR of scabies showed waves of phase-wise increases and decreases throughout the 30 years. The largest increase was seen from 2010 to 2015 with a peak occurring in 2015 (Supplementary Figure 2). The ASIR is approximately 3 times higher than the ASPR, suggesting that, on average, the duration of disease is 4 months or a third of a year (since prevalence rate ≅ incidence rate * duration of disease).

### Age-sex differences in scabies

Males had a slightly higher burden of scabies compared to females as their ASIR and ASPR at the global level decreased from 8.43 (95% CI: 7.47 to 9.48) in 1990 to 8.16 (95% CI: 7.25 to 9.14) in 2021 and from 2.80 (95% CI: 2.48 to 3.14) to 2.70 (95% CI: 2.39 to 3.03), respectively (Supplementary Figure 3). For women, ASIR and ASPR decreased from 8.11 (95% CI: 7.18 to 9.13) in 1990 to 7.94 (95% CI: 7.07 to 8.92) in 2021 and from 2.68 (95% CI: 2.38 to 3.03) in 1990 to 2.63 (95% CI: 2.34 to 2.96) in 2021, respectively.

The gender difference in scabies rates was more pronounced in high and high-middle SDI regions (Supplementary Figure 3). Scabies incidence and prevalence counts show an increasing trend until the 5-to-9-year age group peak, and then gradually decreases (Supplementary Figure 4 (A), (C)). The rates however peak at the 2-to-4-year age group and decrease thereafter, before starting to rise again beyond the 60-year age mark (Supplementary Figure 4 (B), (D)).

### Scabies in major GBD regions

Oceania (ASIR_2021_ = 23.78; 95% CI 21.29 to 26.56; ASPR_2021_ = 7.89; 95% CI 7.02 to 8.89) had the highest ASPR and ASIR per 100 individuals, followed by Tropical Latin America (ASIR_2021_ = 17.92; 95% CI 16.01 to 20.29; ASPR_2021_ = 5.96; 95% CI 5.31 to 6.77) and East Asia (ASIR_2021_ = 14.56; 95% CI 21.29 to 26.56; ASPR_2021_ = 4.81; 95% CI 4.28 to 5.40) in both 1990 and 2019 (Table 1). There were no significant reductions in prevalence or incidence of scabies over the last thirty years in these regions. Age-standardized rates have been the highest in middle SDI regions (ASIR_2021_ = 11.41; 95% CI 10.17 to 12.80; ASPR_2021_ = 3.77; 95% CI 3.36 to 4.22), followed by low-middle and high-middle SDI. Similarly, upper-middle income regions showed the highest rates of incidence and prevalence both in 1990 and 2021, followed by lower-middle- and low-income regions.

### National level divide in scabies

As of 2021, Fiji had the highest age standardized rates of scabies incidence and prevalence (ASIR = 27.17, 95% CI 23.45 to 29.37; ASPR = 8.77, 95% CI 7.75 to 9.83). Fiji was closely followed by other Pacific countries: Papua New Guinea (ASIR = 23.67, 95% CI 21.16 to 26.49; ASPR = 7.86, 95% CI 6.98 to 8.83) the Solomon Islands (ASIR = 23.65, 95% CI 21.10 to 26.54; ASPR = 7.85, 95% CI 6.98 to 8.83) (Figure 1). This trend has been consistent since 1990.

Over the past 30 years, scabies prevalence and incidence rates have significantly increased in forty-six and forty-seven countries respectively, while a significant decline has occurred in forty-nine and fifty-one countries, respectively (Supplementary Table 1). These changes, although statistically significant, were small.

**Figure 1:**
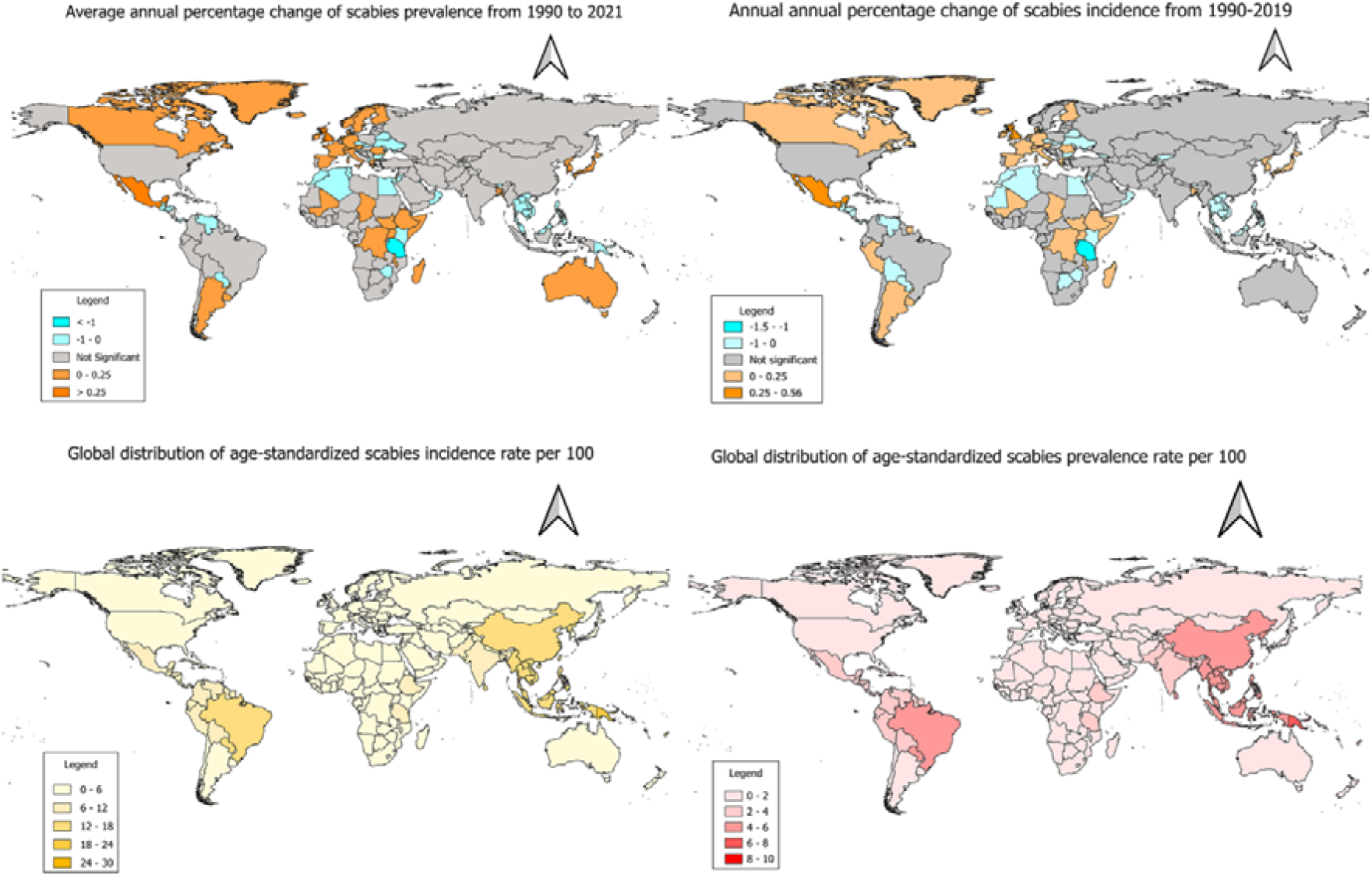
Trends of scabies incidence and prevalence globally

The steepest decline in ASIR from 1990 to 2021 was seen in mostly African countries: Tanzania (AAPC =-1.50; *p*<0.001), Egypt (AAPC = -0.54; *p<*0.001), and Kenya (AAPC = - 0.29; *p<*0.001); and some Pacific and Asian countries: Vanuatu (AAPC = -0.05; *p<*0.001), Lao (AAPC = -0.02; *p<*0.001) and Vietnam (AAPC = -0.01; *p<*0.01). In contrast, the biggest rise was seen in Mexico (AAPC = 0.56; *p<*0.001) followed by the United Kingdom (AAPC = 0.27; *p<*0.001), Romania (AAPC = 0.14; *p<*0.001) and Italy (AAPC = 0.12; *p =* 0.01).

### Temporal trends in scabies

The temporal trend of scabies infestation in the major GBD regions show some surprising insights (Figure 2), with reduced prevalence in Africa, the Middle East and Oceania and increased prevalence in Europe, and the Americas. While a decreasing trend was seen in the sub-Saharan African regions and Oceania (AAPC = -0.02; 95% CI: -0.03 to 0.00), there was a marked increase in the prevalence rates in the Australasian region (AAPC = 0.05; 95% CI: 0.01 to 0.09) and parts of Europe – Central Europe (AAPC = 0.03; 95% CI 0.03 to 0.03) and Eastern Europe (AAPC = 0.11, 95% CI: 0.10 to 0.11). The largest increases occurred in Central Latin America with an AAPC of 0.28 (95% CI: 0.27 to 0.30) followed by Eastern Europe and Australasia. The largest decreases were in Eastern Sub-Saharan Africa with an AAPC of -0.34 (95% CI: -0.29 to -0.34) (Figure 2).

**Figure 2:**
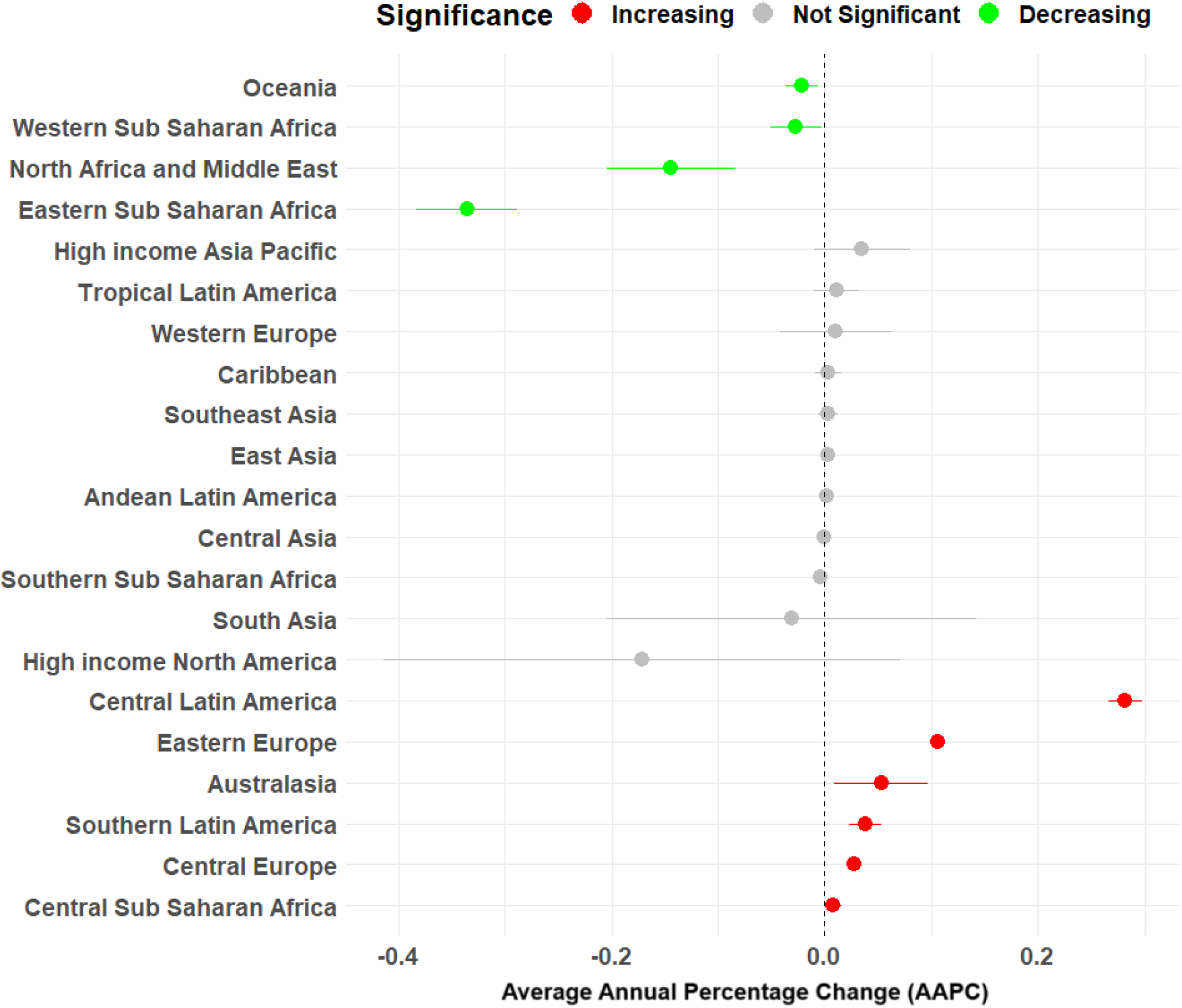
Changes in scabies prevalence in the major global regions

### Relationship between scabies and SDI

The relationship between scabies and SDI is complex, with an ‘n’ shaped relationship between the two variables. Scabies rates have been the highest in middle SDI regions whilst regions on the lower end of the SDI have had scabies rates decline since the 1990s. In contrast, rates have increased in high-middle and high SDI regions, despite their overall low prevalence.

Figure 3 shows an overall stagnant pattern of scabies rates in most other major regions except for Oceania, Tropical Latin America, High-income North America and South-east Asia where rates have increased halfway through the study period before recently returning to their previous level. The moving average (LOWESS) curve shown by the black line indicates a pattern where rates increase from low to middle SDI values before reducing as SDI increases further.

**Figure 3:**
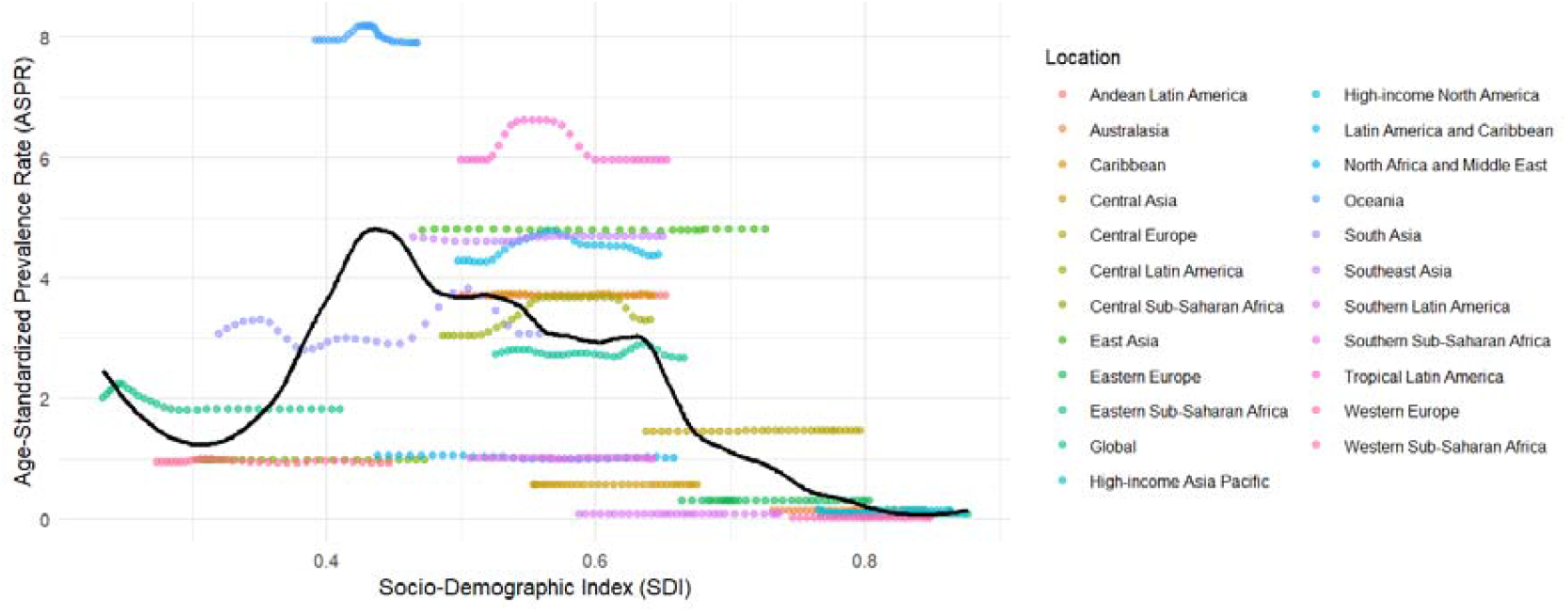
LOWESS curve showing association between SDI and ASPR

### Spatial Clustering

Moran’s *I* for global distribution of scabies rates was 0.78 indicating a high degree of spatial clustering (Supplementary Figure 5). Scabies ‘hotspots’, or clustering of high prevalence areas, were detected in Tropical Latin America, Southeast Asia and Oceania using the bivariate LISA cluster map (**Figure 4**).

**Figure 4:**
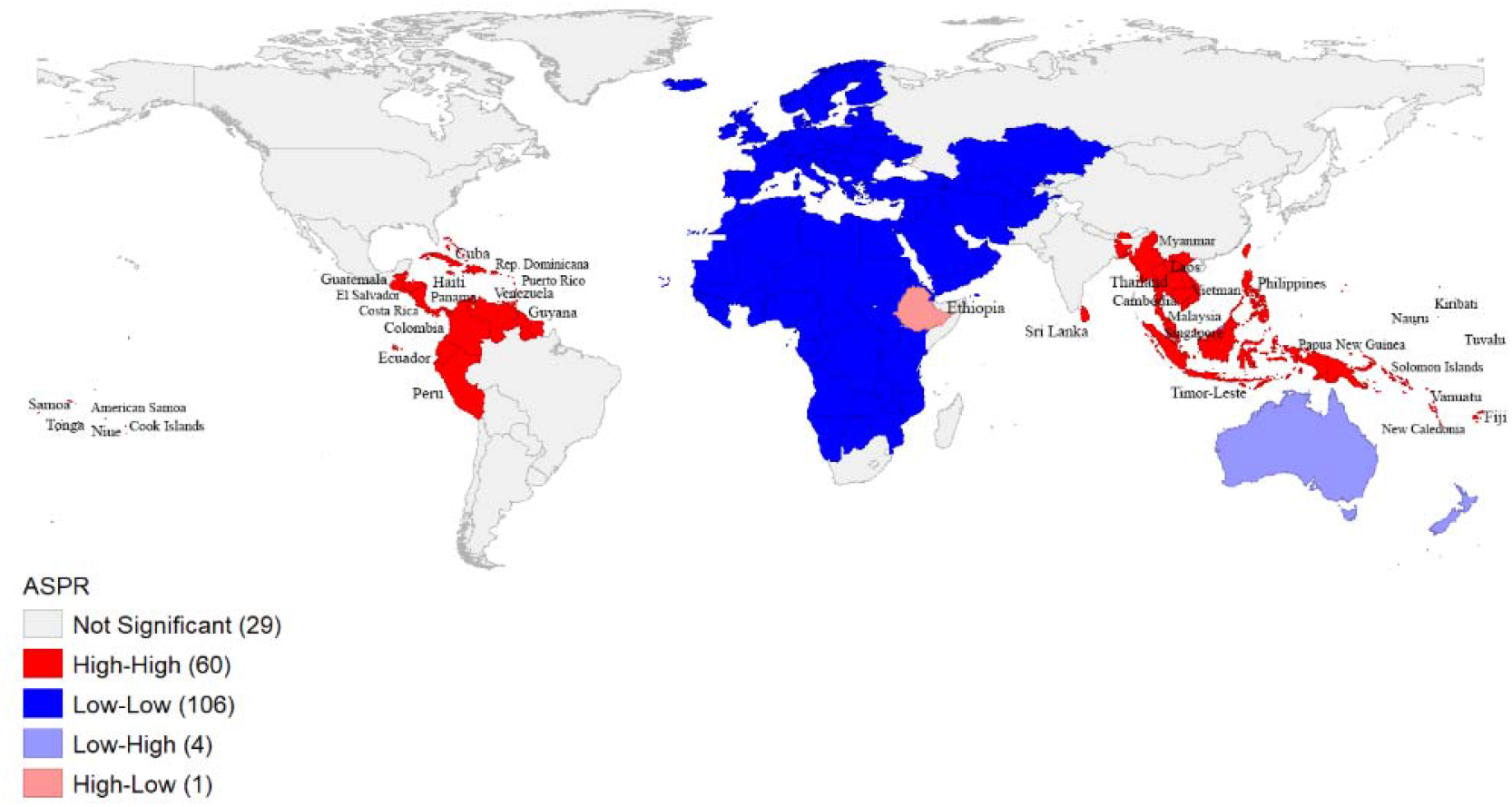
LISA map showing scabies clustering in high and low risk regions

### Risk Factors

As shown in Supplementary table 2, latitude and percentage of urban population were negatively associated with scabies frequency in a log-linear model. While scabies rates decreased on average by 5.52% with each one-degree change in latitude away from the equator, disease frequency approximately declined by 2.74% with each one percentage point increase in urbanization of the country’s population. No significant association was found between population density, percentage of unemployment in the population, life expectancy at birth, gross national income and scabies frequency in the multi-variable model. A summary of model performance has been shown in Supplementary Figure 6.

## Discussion

In this analysis of GBD data from 21 regions including 204 countries or territories from 1990 to 2021 we have found a high burden of scabies clustered in Oceania, Tropical Latin America and Southeast Asia. These findings are consistent with previous GBD estimates (15,16) and other prevalence studies (17-19) that have reported high prevalence in countries near the tropics. The highest prevalence of scabies was observed in Pacific countries with example being Fiji, Papua New Guinea and Solomon Islands. Our study has shown periodic fluctuation in global scabies incidence and prevalence which is very different from the last GBD study that estimated a consistent downward trend of scabies from 1990 to 2017 (15). This is probably due to the recent change in modelling strategies and addition of new data sources (9), that may have changed estimates. Consistent with other studies, the data reported here show higher age-standardized rates of scabies in the young and elderly population (15).

This study has several limitations. Since the inferences of this study is based on the GBD estimates, it shares the shortcomings of this method for estimating disease burden. The GBD data does not consider differences in prevalence rates due to differences in diagnostic techniques (20). Although the GBD estimates improve our understanding of the disease burden, especially for a disease like scabies where prevalence information is limited, it fails to account for underreporting.

Underreporting of scabies is a major problem especially in resource poor nations where patients may not seek healthcare due to lack of knowledge about the disease (21), possible stigmatization and economic constraints. This is why there has been noticeable differences in prevalence rates between national surveys in countries like Fiji and the Solomon Islands (22), reporting a prevalence of over 10%, and the GBD estimates. This means that true prevalence is likely underestimated.

Thus, after adjusting for this underreporting, focus prevalence studies should be conducted in areas with prevalence ranging from 5 to 10% identified by these GBD estimates and mass-drug administration (MDA) to treat scabies must be carried out in regions with high scabies infestation.

Due to the lack of an objective tests for scabies, it is often misdiagnosed, even in high-income countries like New Zealand, which may in turn result in underestimation of the actual burden (23). The GBD considers imputing data from neighbouring countries and predictive covariates in case of missing data which would likely distort the estimates.

While this study provides an overview of scabies prevalence at the global and national levels, it is important to appreciate subnational differences. Since the WHO recommends MDA in regions with prevalence higher than 10% (24), it is essential to identify these pockets of high prevalence even in countries with an otherwise low prevalence.

For example, although overall prevalence of scabies is low overall in Australia, New Zealand or certain parts of Europe there have been reported instances where the prevalence is over 10% in certain communities or regions within these countries. Scabies is endemic in Aboriginal communities in northern Australia (25). Scabies prevalence is high in early childcare centres in socio-economically challenged areas of Auckland, New Zealand (26). Even in Europe, where prevalence of scabies is very low, outbreaks are seen in settings like care homes for the elderly, refugee camps and hospitals in the United Kingdom (27), Greece(28)and Italy (29). Since scabies can potentially lead to further complications, these pockets of high prevalence must be taken seriously, with prevalence documented and MDA considered.

The stagnant level of scabies rates reductions across the major regions of the world is concerning. Although there has been no alarming increase in the scabies rates, the consistent level of high prevalence in Oceania, Tropical Latin America and Southeast Asia indicates lack of progress in disease control in the last three decades. A meta-analysis of MDA trials has found a relative reduction of scabies prevalence by 79% can be achieved with appropriate interventions (7). Although, MDA has started in some Pacific nations like Fiji and the Solomon Islands, there is an urgent need to prioritize interventions in all regions with a prevalence greater than 10%.

The study has revealed new information regarding potential drivers of scabies prevalence. First, latitude and urbanization were significant factors associated with scabies. No other study has considered the influence of latitude as a risk factor for scabies although it is often referred to as “a tropical disease”. Our results are intuitive in a sense that countries farther away from the equator or cooler places have lower scabies rates than warmer counterparts. Increased urban populations may lead to poor living conditions, overcrowding and poor housing which might explain increases urbanisation being associated with high scabies rates. A null association with income levels and scabies differed from previous prevalence studies that found a higher risk of scabies in low-income households (30,31). This may be due to our study’s control of a range of confounders. As scabies is likely to be influenced by geographical factors like precipitation, humidity, elevation and temperature (32), further research on understanding the association of these factors is recommended.

To better estimate the true burden of scabies, it is important to reliably and consistently detect cases. Recent developments, such as the development of objective tests like qPCR and dPCR taken from swabs of skin lesions may help. The International Alliance for Control of Scabies (IACS) consensus criteria also presents a robust set of diagnostic criteria which promises to improve sensitivity of scabies detection in a clinical context (33).

Although IACS has been used in some prevalence studies, its global adoption is lacking. Our study indicates that global scabies prevalence remains high and progress to reduce scabies prevalence is slow with existing programs. Scabies control should be further prioritized to accelerate progress to reduce the prevalence of this important neglected tropical disease. Finally, inspiration should be drawn from programmes conducted in Pacific Island nations such as Fiji and the Solomon Islands where nationwide MDA has been used to significantly reduce the prevalence and complications of scabies.

## Data Availability

All data produced are available online at: http://www.healthdata.org/gbd/ and https://data.worldbank.org

https://github.com/Saptorshi1999/Scabies_code_GBD_Study.git

## Data Sharing

Data for this study has been extracted from the Global Burden of Disease (GBD) study, 2021: https://vizhub.healthdata.org/gbd-results/

## Code

Codes for statistical analysis of this paper can be found at: https://github.com/Saptorshi1999/Scabies_code_GBD_Study.git

## Author Roles

SG Conceptualization, Data Curation, Formal analysis, Methodology, Software, Validation, Visualization, Writing - original draft, review & editing. **ST:** Conceptualization, Data Curation, Formal analysis, Funding acquisition, Investigation, Project administration, Resources, Software, Supervision, Validation, Writing - review & editing. **AM:** Validation, Writing - review & editing. **GS:** Funding acquisition, Project administration, Resources, Supervision, Validation, Writing - review & editing. **CG:** Validation, Writing - review & editing.

## Notes

### Competing Interest Statement

The authors have declared no competing interest.

### Funding Statement

This project is funded by the Health Research Council of New Zealand (grant number: 22/269).

### Author Declarations

The data can be accessed from: https://vizhub.healthdata.org/gbd-results/

## References

1. Heukelbach, J., & Feldmeier, H. (2006). Scabies. Lancet (London, England), 367(9524), 1767–1774. 10.1016/S0140-6736(06)68772-2

2. Tong, S. Y., Davis, J. S., Eichenberger, E., Holland, T. L., & Fowler, V. G., Jr (2015). Staphylococcus aureus infections: epidemiology, pathophysiology, clinical manifestations, and management. Clinical microbiology reviews, 28(3), 603–661. 10.1128/CMR.00134-14

3. Chisavu, F., Gafencu, M., Steflea, R. M., Vaduva, A., Izvernariu, F., & Stroescu, R. F. (2024). Acute Glomerulonephritis Following Systemic Scabies in Two Brothers. Children, 11(8), 981. 10.3390/children11080981

4. Engelman, D., Kiang, K., Chosidow, O., McCarthy, J., Fuller, C., Lammie, P., Hay, R., et al (2013). Toward the global control of human scabies: introducing the International Alliance for the Control of Scabies. PLoS neglected tropical diseases, 7(8), e2167. 10.1371/journal.pntd.0002167

5. Parks, T., Smeesters, P. R., & Steer, A. C. (2012). Streptococcal skin infection and rheumatic heart disease. Current opinion in infectious diseases, 25(2), 145–153. 10.1097/QCO.0b013e3283511d27

6. Fernando, D. D., Mounsey, K. E., Bernigaud, C., Surve, N., Estrada Chávez, G. E., Hay, R. J., Currie, B. J., et al (2024). Scabies. Nature reviews. Disease primers, 10(1), 74. 10.1038/s41572-024-00552-8

7. Lake, S. J., Kaldor, J. M., Hardy, M., Engelman, D., Steer, A. C., & Romani, L. (2022). Mass Drug Administration for the Control of Scabies: A Systematic Review and Meta-analysis. Clinical infectious diseases : an official publication of the Infectious Diseases Society of America, 75(6), 959–967. 10.1093/cid/ciac042

8. Global Burden of Disease Study 2021 (GBD 2021) Socio-Demographic Index (SDI) 1950–2021 | GHDx [Internet]. [cited 2024 Oct 6]. Available from: https://ghdx.healthdata.org/record/global-burden-disease-study-2021-gbd-2021-socio-demographic-index-sdi-1950%E2%80%932021

9. GBD 2021 Diseases and Injuries Collaborators (2024). Global incidence, prevalence, years lived with disability (YLDs), disability-adjusted life-years (DALYs), and healthy life expectancy (HALE) for 371 diseases and injuries in 204 countries and territories and 811 subnational locations, 1990-2021: a systematic analysis for the Global Burden of Disease Study 2021. Lancet (London, England), 403(10440), 2133–2161. 10.1016/S0140-6736(24)00757-8

10. Scabies | Institute for Health Metrics and Evaluation [Internet]. [cited 2024 Oct 6]. Available from: https://www.healthdata.org/gbd/methods-appendices-2021/scabies

11. World Bank Open Data [Internet]. [cited 2024 Aug 8]. World Bank Open Data. Available from: https://data.worldbank.org

12. R: The R Project for Statistical Computing [Internet]. [cited 2024 Aug 8]. Available from: https://www.r-project.org/

13. QGIS overview · QGIS Web Site [Internet]. [cited 2024 Sep 25]. Available from: https://www.qgis.org/project/overview/

14. Joinpoint Help System [Internet]. [cited 2024 Oct 6]. Citation. Available from: https://surveillance.cancer.gov/help/joinpoint/tech-help/citation

15. Zhang, W., Zhang, Y., Luo, L., Huang, W., Shen, X., Dong, X., Zeng, W., & Lu, H. (2020). Trends in prevalence and incidence of scabies from 1990 to 2017: findings from the global Burden of disease study 2017. Emerging microbes & infections, 9(1), 813–816. 10.1080/22221751.2020.1754136

16. Karimkhani, C., Colombara, D. V., Drucker, A. M., Norton, S. A., Hay, R., Engelman, D., Steer, A., Whitfeld, M., Naghavi, M., & Dellavalle, R. P. (2017). The global burden of scabies: a cross-sectional analysis from the Global Burden of Disease Study 2015. The Lancet. Infectious diseases, 17(12), 1247– 1254. 10.1016/S1473-3099(17)30483-8

17. Schneider, S., Wu, J., Tizek, L., Ziehfreund, S., & Zink, A. (2023). Prevalence of scabies worldwide-An updated systematic literature review in 2022. Journal of the European Academy of Dermatology and Venereology : JEADV, 37(9), 1749–1757. 10.1111/jdv.19167

18. Romani, L., Steer, A. C., Whitfeld, M. J., & Kaldor, J. M. (2015). Prevalence of scabies and impetigo worldwide: a systematic review. The Lancet. Infectious diseases, 15(8), 960–967. 10.1016/S1473-3099(15)00132-2

19. Aždajić, M. D., Bešlić, I., Gašić, A., Ferara, N., Pedić, L., & Lugović-Mihić, L. (2022). Increased Scabies Incidence at the Beginning of the 21st Century: What Do Reports from Europe and the World Show?. Life (Basel, Switzerland), 12(10), 1598. 10.3390/life12101598

20. Gupta, S., Thornley, S., Morris, A., Sundborn, G., & Grant, C. (2024). Prevalence and determinants of scabies: A global systematic review and meta-analysis. Tropical medicine & international health : TM & IH, 29(12), 1006–1017. 10.1111/tmi.14058

21. Lopes, M. J., da Silva, E. T., Ca, J., Gonçalves, A., Rodrigues, A., Mandjuba, C., Nakutum, J., et al. (2020). Perceptions, attitudes and practices towards scabies in communities on the Bijagós Islands, Guinea-Bissau. Transactions of the Royal Society of Tropical Medicine and Hygiene, 114(1), 49–56. 10.1093/trstmh/trz102

22. Tsoi, S. K., Lake, S. J., Thean, L. J., Matthews, A., Sokana, O., Kama, M., Amaral, S., et al. (2021). Estimation of scabies prevalence using simplified criteria and mapping procedures in three Pacific and southeast Asian countries. BMC public health, 21(1), 2060. 10.1186/s12889-021-12039-2

23. Thornley, S., Sundborn, G., Engelman, D., Roskvist, R., Pasay, C., Marshall, R., Long, W., et al. (2023). Children’s scabies survey indicates high prevalence and misdiagnosis in Auckland educational institutions. Journal of paediatrics and child health, 59(12), 1296–1303. 10.1111/jpc.16512

24. Engelman D, Marks M, Steer AC, Beshah A, Biswas G, Chosidow O, et al. A framework for scabies control. Taylan Ozkan A, editor. PLoS Negl Trop Dis. 2021 Sep 2;15(9):e0009661.

25. Gramp, P., & Gramp, D. (2021). Scabies in remote Aboriginal and Torres Strait Islander populations in Australia: A narrative review. PLoS neglected tropical diseases, 15(9), e0009751. 10.1371/journal.pntd.0009751

26. Thornley, S., Sundborn, G., Engelman, D., Roskvist, R., Heather, M., Pasay, C. J., Marshall, R., & McCarthy, J. (2022). High prevalence of scabies in Auckland pre-schools. The New Zealand medical journal, 135(1560), 12–17. 10.26635/6965.5610

27. Cassell, J. A., Middleton, J., Nalabanda, A., Lanza, S., Head, M. G., Bostock, J., Hewitt, K., Jones, C. I., Darley, C., Karir, S., & Walker, S. L. (2018). Scabies outbreaks in ten care homes for elderly people: a prospective study of clinical features, epidemiology, and treatment outcomes. The Lancet. Infectious diseases, 18(8), 894–902. 10.1016/S1473-3099(18)30347-5

28. Louka, C., Logothetis, E., Engelman, D., Samiotaki-Logotheti, E., Pournaras, S., & Stienstra, Y. (2022). Scabies epidemiology in health care centers for refugees and asylum seekers in Greece. PLoS neglected tropical diseases, 16(6), e0010153. 10.1371/journal.pntd.0010153

29. Sponselli, S., De Maria, L., Caputi, A., Stefanizzi, P., Bianchi, F. P., Delvecchio, G., Foti, C., Romita, P., Ambrogio, F., Zagaria, S., Giannelli, G., Tafuri, S., & Vimercati, L. (2023). Infection Control among Healthcare Workers and Management of a Scabies Outbreak in a Large Italian University Hospital. Journal of clinical medicine, 12(11), 3830. 10.3390/jcm12113830

30. Ugbomoiko, U. S., Oyedeji, S. A., Babamale, O. A., & Heukelbach, J. (2018). Scabies in Resource-Poor Communities in Nasarawa State, Nigeria: Epidemiology, Clinical Features and Factors Associated with Infestation. Tropical medicine and infectious disease, 3(2), 59. 10.3390/tropicalmed3020059

31. Ejigu, K., Haji, Y., Toma, A., & Tadesse, B. T. (2019). Factors associated with scabies outbreaks in primary schools in Ethiopia: a case-control study. Research and reports in tropical medicine, 10, 119– 127. 10.2147/RRTM.S214724

32. Liu, J. M., Wang, H. W., Chang, F. W., Liu, Y. P., Chiu, F. H., Lin, Y. C., Cheng, K. C., & Hsu, R. J. (2016). The effects of climate factors on scabies. A 14-year population-based study in Taiwan. Les effets des facteurs climatiques sur la gale. Une étude de 14 ans sur la population à Taiwan. Parasite (Paris, France), 23, 54. 10.1051/parasite/2016065

33. Engelman, D., Yoshizumi, J., Hay, R. J., Osti, M., Micali, G., Norton, S., Walton, S., et al. (2020). The 2020 International Alliance for the Control of Scabies Consensus Criteria for the Diagnosis of Scabies. The British journal of dermatology, 183(5), 808–820. 10.1111/bjd.18943

