## Supplementary File for "Global trends and prevalence of scabies: a spatiotemporal analysis using Global Burden of Disease 2021 data"

**Supplementary Material**

Contents

| **Sl. No.** | **Topic** | **Page No.** |
| --- | --- | --- |
| 1 | Supplementary Figure 1: Conceptual Framework for data collection and analysis | **2** |
| 2 | Supplementary Figure 2: Global age-standardized incidence and prevalence rates of scabies from 1990 to 2021 | **3** |
| 3 | Supplementary Figure 3: Gender differences in age standardized incidence and prevalence rates of scabies by SDI regions | **4** |
| 4 | Supplementary Figure 4: Age-sex distribution of scabies cases globally | **5** |
| 5 | Supplementary Table 1: Trends in age-standardized scabies prevalence and incidence rates for countries | **6-10** |
| 6 | Supplementary Figure 5: Spatial Clustering and Moran’s *I* | **11** |
| 7 | Supplementary Table 2: Association of scabies rates with socioeconomic factors | **12** |
| 8 | Supplementary Figure 6: Model diagnostics | **13** |

**Supplementary Figure 1: Conceptual Framework for data collection and analysis**

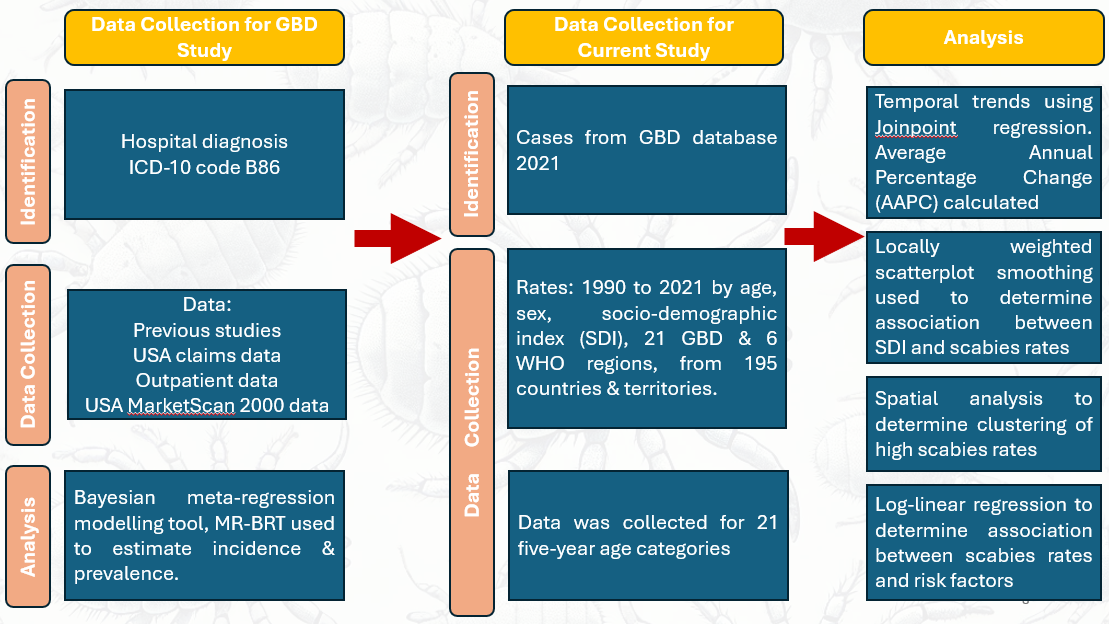

**Supplementary Figure 2: Global age-standardized incidence and prevalence rates of scabies from 1990 to 2021**

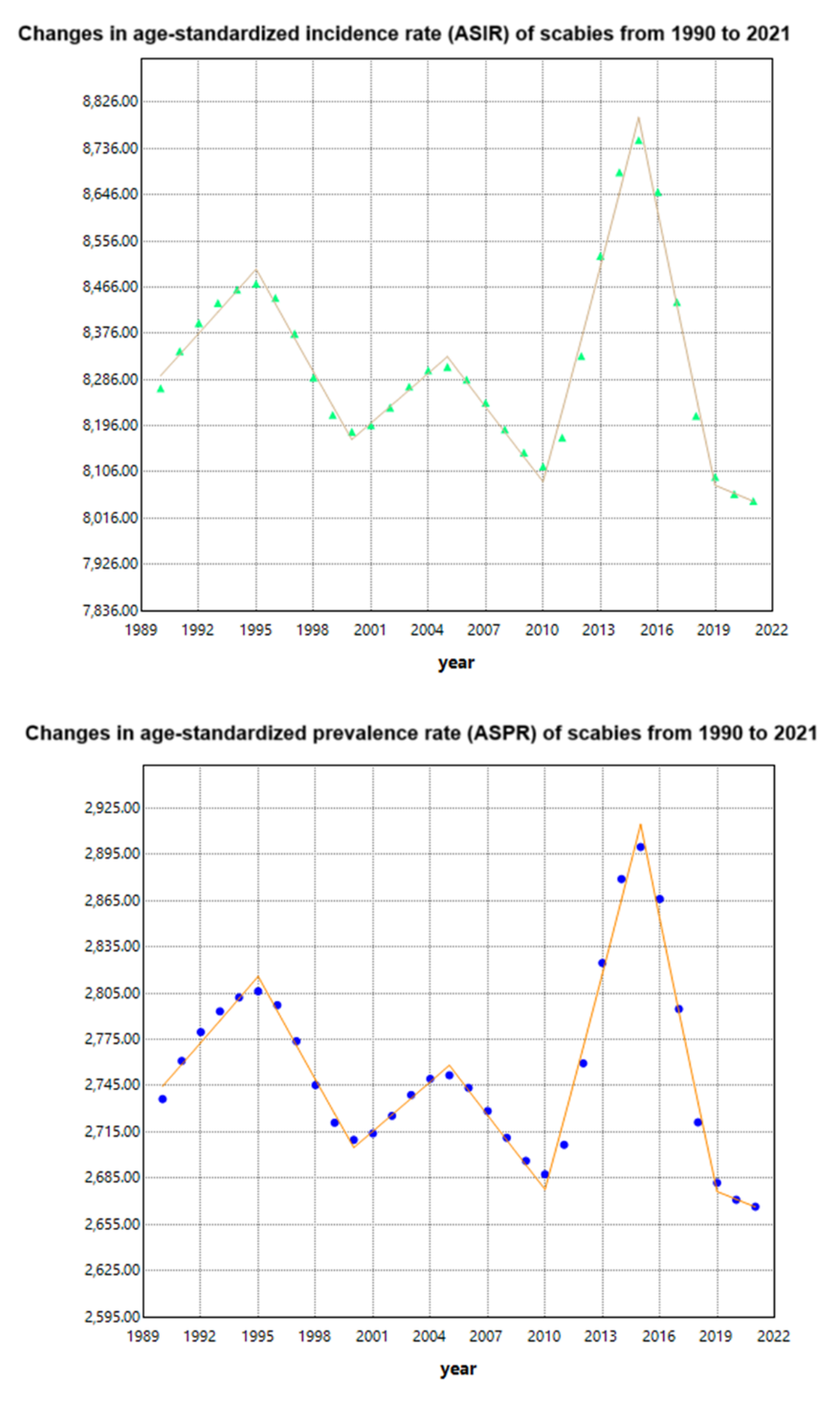

**Supplementary Figure 3: Gender differences in age standardized incidence and prevalence rates of scabies by SDI regions**

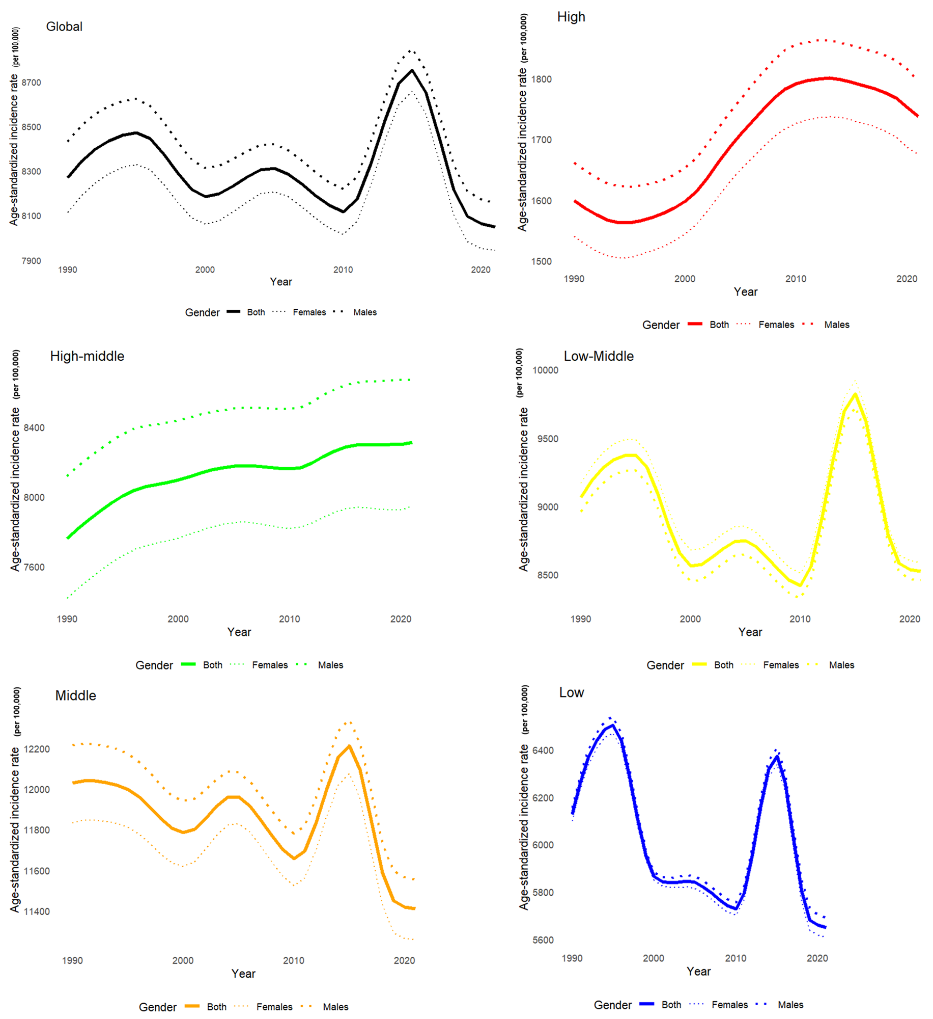

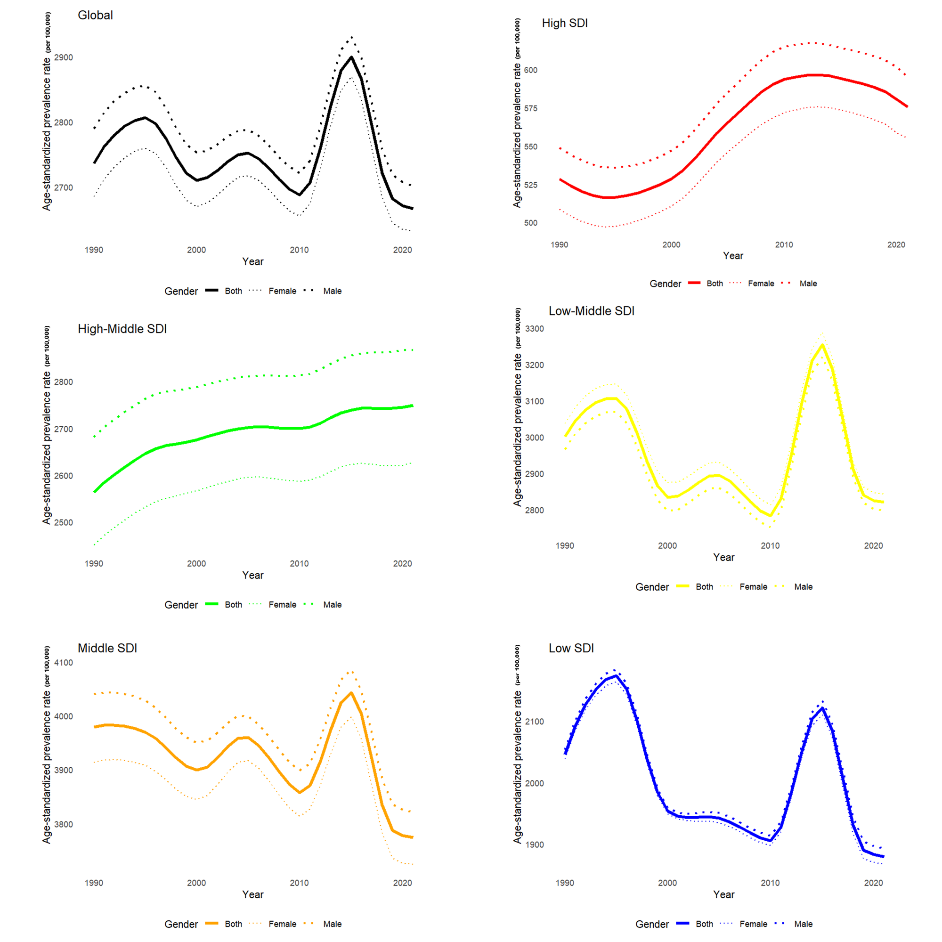

**Supplementary Figure 4: Age-sex distribution of scabies cases globally**

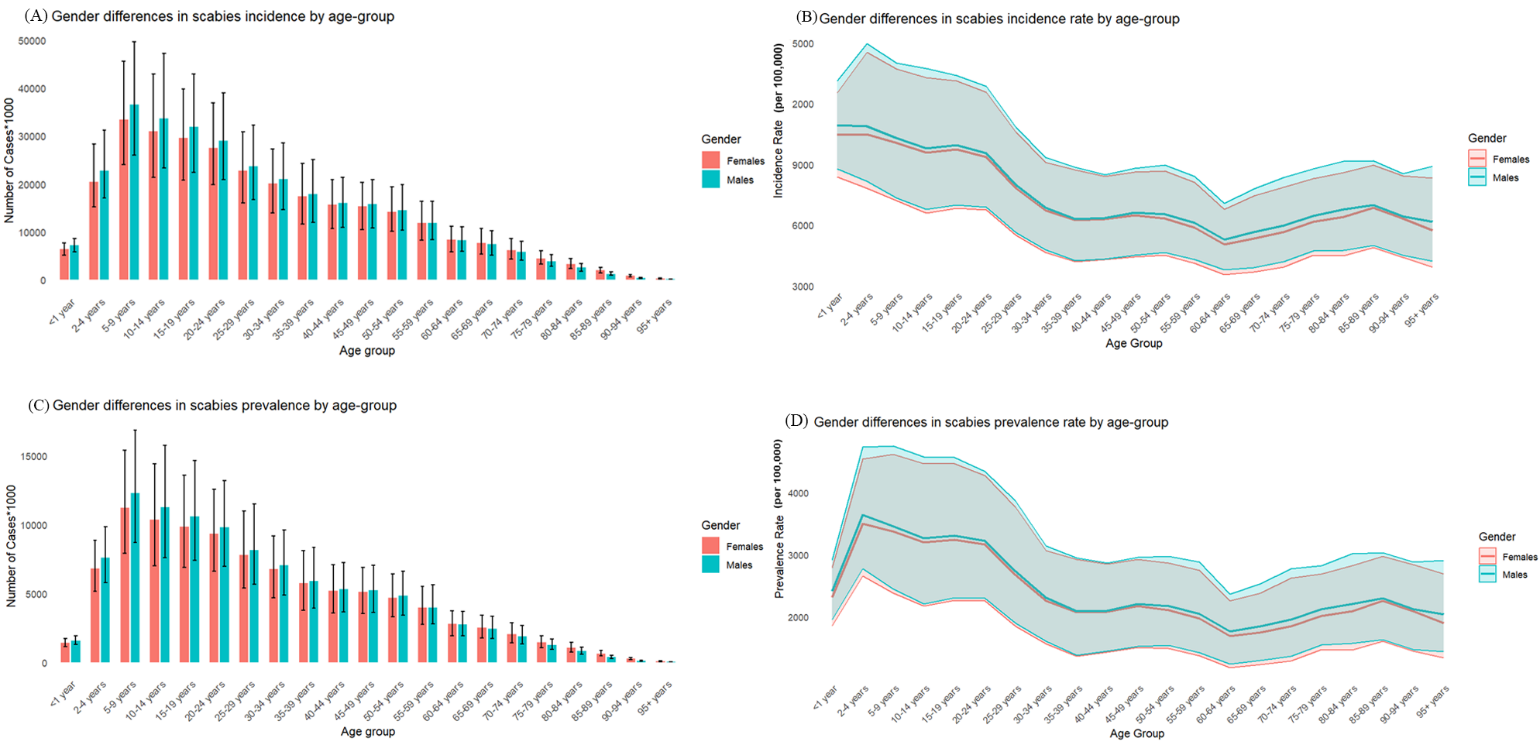

**Supplementary Table 1: Trends in age-standardized scabies prevalence and incidence rates for countries**

| **location_name** | **ASPR (per 100)** | **AAPC** | **P-Value** | **location_name** | **ASIR (per 100)** | **AAPC** | **P-Value** |
| --- | --- | --- | --- | --- | --- | --- | --- |
| ***Decreasing trend*** | | | | | | | |
| United Republic of Tanzania | 2.20 | -1.613 | <0.001 | United Republic of Tanzania | 6.37 | -1.502 | <0.001 |
| Egypt | 1.29 | -0.581 | <0.001 | Egypt | 3.83 | -0.538 | <0.001 |
| Kenya | 1.87 | -0.304 | <0.001 | Kenya | 5.63 | -0.294 | <0.001 |
| Vanuatu | 6.99 | -0.055 | <0.001 | Vanuatu | 21.27 | -0.052 | <0.001 |
| Lao People's Democratic Republic | 4.54 | -0.016 | <0.001 | Lao People's Democratic Republic | 13.73 | -0.016 | <0.001 |
| Vietnam | 4.53 | -0.012 | <0.01 | Vietnam | 13.72 | -0.013 | <0.05 |
| Morocco | 0.96 | -0.012 | <0.001 | Gambia | 1.88 | -0.012 | <0.01 |
| Maldives | 4.54 | -0.011 | <0.001 | Maldives | 13.73 | -0.012 | <0.001 |
| Oman | 0.96 | -0.011 | <0.001 | Morocco | 2.91 | -0.011 | <0.001 |
| Guatemala | 3.20 | -0.010 | <0.001 | Thailand | 13.74 | -0.011 | <0.001 |
| Palestine | 0.96 | -0.010 | <0.001 | Guatemala | 9.67 | -0.010 | <0.001 |
| Thailand | 4.54 | -0.010 | <0.001 | Oman | 2.91 | -0.010 | <0.001 |
| Gambia | 0.62 | -0.009 | <0.001 | Czech Republic | 4.32 | -0.009 | <0.001 |
| El Salvador | 3.20 | -0.009 | <0.001 | Palestine | 2.91 | -0.009 | <0.001 |
| Nauru | 7.83 | -0.009 | <0.001 | Slovakia | 4.32 | -0.009 | <0.001 |
| Slovakia | 1.44 | -0.008 | <0.001 | Nauru | 23.60 | -0.009 | <0.01 |
| Venezuela | 3.20 | -0.008 | <0.001 | Venezuela | 9.66 | -0.008 | <0.001 |
| Tunisia | 0.96 | -0.008 | <0.001 | Kuwait | 2.91 | -0.008 | <0.001 |
| Czech Republic | 1.44 | -0.008 | <0.001 | Mauritius | 13.70 | -0.008 | <0.001 |
| Kuwait | 0.96 | -0.008 | <0.001 | Marshall Islands | 23.61 | -0.008 | <0.001 |
| Micronesia (Federated States of) | 7.83 | -0.008 | <0.001 | El Salvador | 9.67 | -0.008 | <0.001 |
| Mauritius | 4.53 | -0.007 | <0.001 | Tunisia | 2.91 | -0.008 | <0.001 |
| Marshall Islands | 7.84 | -0.007 | <0.001 | Mauritania | 1.87 | -0.007 | <0.05 |
| Bhutan | 2.96 | -0.007 | <0.001 | Micronesia (Federated States of) | 23.60 | -0.007 | <0.001 |
| Seychelles | 4.52 | -0.007 | <0.001 | Seychelles | 13.69 | -0.007 | <0.001 |
| Philippines | 4.83 | -0.007 | <0.001 | Botswana | 2.92 | -0.007 | <0.01 |
| Cambodia | 4.54 | -0.006 | <0.001 | Philippines | 14.60 | -0.007 | <0.001 |
| Paraguay | 5.70 | -0.005 | <0.001 | Cambodia | 13.74 | -0.007 | <0.001 |
| Zimbabwe | 0.97 | -0.005 | <0.01 | Nicaragua | 9.68 | -0.006 | <0.001 |
| Algeria | 0.96 | -0.005 | <0.001 | Zimbabwe | 2.92 | -0.006 | <0.001 |
| Malaysia | 4.53 | -0.005 | <0.01 | Bahrain | 2.91 | -0.006 | <0.01 |
| Djibouti | 1.37 | -0.005 | <0.001 | Bhutan | 8.92 | -0.006 | <0.001 |
| Slovenia | 1.44 | -0.005 | <0.05 | Cook Islands | 23.58 | -0.005 | <0.001 |
| Jordan | 0.96 | -0.005 | <0.001 | Malaysia | 13.71 | -0.005 | <0.05 |
| Cook Islands | 7.83 | -0.005 | <0.001 | Ukraine | 0.43 | -0.005 | <0.001 |
| Lebanon | 0.96 | -0.005 | <0.001 | Lebanon | 2.91 | -0.005 | <0.001 |
| Bahrain | 0.96 | -0.005 | <0.001 | Paraguay | 16.98 | -0.004 | <0.01 |
| Solomon Islands | 7.85 | -0.004 | <0.001 | Jordan | 2.91 | -0.004 | <0.001 |
| Honduras | 3.20 | -0.004 | <0.05 | Algeria | 2.91 | -0.004 | <0.01 |
| Bulgaria | 1.44 | -0.004 | <0.001 | Djibouti | 4.14 | -0.004 | <0.001 |
| Panama | 3.20 | -0.004 | <0.01 | Solomon Islands | 23.65 | -0.004 | <0.001 |
| Tuvalu | 7.84 | -0.004 | <0.05 | Honduras | 9.67 | -0.004 | <0.05 |
| Belarus | 0.13 | -0.004 | <0.01 | Bulgaria | 4.33 | -0.004 | <0.001 |
| Ukraine | 0.14 | -0.004 | <0.05 | Georgia | 1.74 | -0.003 | <0.05 |
| Papua New Guinea | 7.86 | -0.003 | <0.001 | Panama | 9.67 | -0.003 | <0.01 |
| Albania | 1.45 | -0.002 | <0.05 | Burundi | 4.15 | -0.003 | <0.001 |
| Burundi | 1.38 | -0.002 | <0.05 | Bolivia | 11.19 | -0.002 | <0.01 |
| Samoa | 7.83 | -0.001 | <0.05 | Belarus | 0.40 | -0.002 | <0.05 |
| Tonga | 7.84 | -0.001 | <0.05 | Kyrgyzstan | 1.74 | -0.002 | <0.05 |
|  |  |  |  | Bahamas | 11.18 | -0.002 | <0.05 |
|  |  |  |  | Samoa | 23.59 | -0.002 | <0.05 |
| ***Not Significant*** | | | | | | | |
| Botswana | 0.96 | Not Significant | | Slovenia | 4.32 | Not significant | |
| Russian Federation | 0.38 | Not Significant | | Tuvalu | 23.61 | Not significant | |
| Liberia | 0.62 | Not Significant | | Papua New Guinea | 23.67 | Not significant | |
| Suriname | 3.72 | Not Significant | | Albania | 4.34 | Not significant | |
| Nigeria | 1.32 | Not Significant | | Tonga | 23.63 | Not significant | |
| Georgia | 0.58 | Not Significant | | Russian Federation | 1.13 | Not significant | |
| Chile | 0.08 | Not Significant | | Liberia | 1.88 | Not significant | |
| Afghanistan | 0.96 | Not Significant | | Nigeria | 3.95 | Not significant | |
| Eswatini | 0.96 | Not Significant | | Chile | 0.23 | Not significant | |
| Comoros | 1.38 | Not Significant | | Afghanistan | 2.92 | Not significant | |
| Tokelau | 7.83 | Not Significant | | Eswatini | 2.92 | Not significant | |
| Kyrgyzstan | 0.58 | Not Significant | | Comoros | 4.15 | Not significant | |
| Tajikistan | 0.58 | Not Significant | | Tokelau | 23.59 | Not significant | |
| Equatorial Guinea | 0.97 | Not Significant | | Tajikistan | 1.75 | Not significant | |
| Nicaragua | 3.21 | Not Significant | | Equatorial Guinea | 2.95 | Not significant | |
| Indonesia | 4.83 | Not Significant | | Indonesia | 14.59 | Not significant | |
| Portugal | 0.01 | Not Significant | | Portugal | 0.03 | Not significant | |
| Sao Tome and Principe | 0.62 | Not Significant | | Sao Tome and Principe | 1.87 | Not significant | |
| Syrian Arab Republic | 0.96 | Not Significant | | Syrian Arab Republic | 2.91 | Not significant | |
| United States of America | 0.13 | Not Significant | | United States of America | 0.40 | Not significant | |
| Singapore | 0.08 | Not Significant | | Singapore | 0.23 | Not significant | |
| Lesotho | 0.97 | Not Significant | | Lesotho | 2.93 | Not significant | |
| United Arab Emirates | 0.96 | Not Significant | | United Arab Emirates | 2.91 | Not significant | |
| Hungary | 1.44 | Not Significant | | Hungary | 4.32 | Not significant | |
| Sudan | 0.96 | Not Significant | | Sudan | 2.92 | Not significant | |
| Taiwan | 4.53 | Not Significant | | Taiwan | 13.74 | Not significant | |
| Cyprus | 0.01 | Not Significant | | Iraq | 2.21 | Not significant | |
| Iraq | 0.71 | Not Significant | | Poland | 4.59 | Not significant | |
| Poland | 1.52 | Not Significant | | Belgium | 0.02 | Not significant | |
| Belgium | 0.01 | Not Significant | | Northern Mariana Islands | 23.61 | Not significant | |
| Northern Mariana Islands | 7.83 | Not Significant | | Niue | 23.62 | Not significant | |
| Niue | 7.84 | Not Significant | | Mozambique | 4.16 | Not significant | |
| Mozambique | 1.38 | Not Significant | | Yemen | 2.92 | Not significant | |
| Yemen | 0.96 | Not Significant | | Brazil | 17.95 | Not significant | |
| Brazil | 5.97 | Not Significant | | Saint Lucia | 11.22 | Not significant | |
| Dominica | 3.72 | Not Significant | | Côte d'Ivoire | 1.88 | Not significant | |
| Saint Lucia | 3.72 | Not Significant | | Armenia | 1.74 | Not significant | |
| Côte d'Ivoire | 0.62 | Not Significant | | Saint Kitts and Nevis | 11.20 | Not significant | |
| Armenia | 0.58 | Not Significant | | Andorra | 0.03 | Not significant | |
| Saint Kitts and Nevis | 3.72 | Not Significant | | Dominican Republic | 11.22 | Not significant | |
| Andorra | 0.01 | Not Significant | | Congo | 2.95 | Not significant | |
| Mauritania | 0.62 | Not Significant | | Burkina Faso | 1.88 | Not significant | |
| Dominican Republic | 3.72 | Not Significant | | American Samoa | 23.62 | Not significant | |
| Congo | 0.97 | Not Significant | | New Zealand | 0.39 | Not significant | |
| Burkina Faso | 0.62 | Not Significant | | Costa Rica | 9.67 | Not significant | |
| American Samoa | 7.84 | Not Significant | | Cape Verde | 1.87 | Not significant | |
| New Zealand | 0.13 | Not Significant | | Guyana | 11.20 | Not significant | |
| Costa Rica | 3.20 | Not Significant | | Togo | 1.88 | Not significant | |
| Cape Verde | 0.62 | Not Significant | | South Africa | 3.08 | Not significant | |
| Peru | 3.71 | Not Significant | | Cuba | 11.21 | Not significant | |
| Bahamas | 3.71 | Not Significant | | Bosnia & Herzegovina | 4.33 | Not significant | |
| Guyana | 3.72 | Not Significant | | Serbia | 4.33 | Not significant | |
| Togo | 0.62 | Not Significant | | Niger | 1.88 | Not significant | |
| South Africa | 1.02 | Not Significant | | Mongolia | 1.75 | Not significant | |
| Cuba | 3.72 | Not Significant | | Myanmar | 13.75 | Not significant | |
| Bosnia & Herzegovina | 1.44 | Not Significant | | Moldova, Republic of | 0.40 | Not significant | |
| Serbia | 1.44 | Not Significant | | Trinidad and Tobago | 11.18 | Not significant | |
| Niger | 0.62 | Not Significant | | Central African Republic | 2.96 | Not significant | |
| Mongolia | 0.58 | Not Significant | | Turkey | 2.91 | Not significant | |
| Myanmar | 4.55 | Not Significant | | Saudi Arabia | 2.91 | Not significant | |
| Moldova, Republic of | 0.13 | Not Significant | | Barbados | 11.19 | Not significant | |
| Trinidad and Tobago | 3.71 | Not Significant | | Libyan Arab Jamahiriya | 2.91 | Not significant | |
| Central African Republic | 0.98 | Not Significant | | Colombia | 9.67 | Not significant | |
| Turkey | 0.96 | Not Significant | | Iran (Islamic Republic of) | 3.07 | Not significant | |
| Saudi Arabia | 0.96 | Not Significant | | Antigua & Barbuda | 11.19 | Not significant | |
| Barbados | 3.71 | Not Significant | | North Macedonia | 4.33 | Not significant | |
| Libyan Arab Jamahiriya | 0.96 | Not Significant | | Bermuda | 11.19 | Not significant | |
| Colombia | 3.20 | Not Significant | | Uzbekistan | 1.74 | Not significant | |
| Bolivia | 3.71 | Not Significant | | Sierra Leone | 1.88 | Not significant | |
| Iran (Islamic Republic of) | 1.01 | Not Significant | | Ecuador | 11.18 | Not significant | |
| Antigua & Barbuda | 3.71 | Not Significant | | Kiribati | 23.63 | Not significant | |
| North Macedonia | 1.44 | Not Significant | | Qatar | 2.91 | Not significant | |
| Bermuda | 3.71 | Not Significant | | Lithuania | 0.40 | Not significant | |
| Uzbekistan | 0.58 | Not Significant | | Sri Lanka | 12.27 | Not significant | |
| Sierra Leone | 0.62 | Not Significant | | China | 14.59 | Not significant | |
| Ecuador | 3.71 | Not Significant | | India | 9.34 | Not significant | |
| Kiribati | 7.84 | Not Significant | | Jammu-Kashmir | 9.34 | Not significant | |
| Qatar | 0.96 | Not Significant | | Pakistan | 9.40 | Not significant | |
| Lithuania | 0.13 | Not Significant | | Guinea | 1.88 | Not significant | |
| Sri Lanka | 4.01 | Not Significant | | Latvia | 0.40 | Not significant | |
| China | 4.82 | Not Significant | | Croatia | 4.33 | Not significant | |
| India | 3.08 | Not Significant | | Grenada | 11.21 | Not significant | |
| Jammu-Kashmir | 3.08 | Not Significant | | Montenegro | 4.33 | Not significant | |
| Pakistan | 3.11 | Not Significant | | Benin | 1.87 | Not significant | |
| Guinea | 0.62 | Not Significant | | Gabon | 2.95 | Not significant | |
| Latvia | 0.13 | Not Significant | | Eritrea | 4.15 | Not significant | |
| Croatia | 1.44 | Not Significant | | Guinea-Bissau | 1.88 | Not significant | |
| Grenada | 3.72 | Not Significant | | Estonia | 0.16 | Not significant | |
| Montenegro | 1.44 | Not Significant | | Ghana | 0.95 | Not significant | |
| Benin | 0.62 | Not Significant | | Puerto Rico | 11.19 | Not significant | |
| Gabon | 0.97 | Not Significant | | Fiji | 26.17 | Not significant | |
| Eritrea | 1.38 | Not Significant | | Saint Vincent and the Grenadines | 11.22 | Not significant | |
| Guinea-Bissau | 0.62 | Not Significant | | Azerbaijan | 1.75 | Not significant | |
| Estonia | 0.05 | Not Significant | | Haiti | 11.22 | Not significant | |
| Ghana | 0.30 | Not Significant | | Nepal | 8.84 | Not significant | |
| Puerto Rico | 3.71 | Not Significant | | Angola | 2.95 | Not significant | |
| Fiji | 8.77 | Not Significant | | Rwanda | 1.63 | Not significant | |
| Saint Vincent and the Grenadines | 3.72 | Not Significant | | Cameroon | 1.88 | Not significant | |
| Azerbaijan | 0.58 | Not Significant | | Namibia | 2.92 | Not significant | |
| Haiti | 3.72 | Not Significant | | Senegal | 1.87 | Not significant | |
| Nepal | 2.93 | Not Significant | | Turkmenistan | 1.75 | Not significant | |
| Angola | 0.97 | Not Significant | | Timor-Leste | 15.37 | Not significant | |
| Rwanda | 0.51 | Not Significant | | Kazakhstan | 1.74 | Not significant | |
| Cameroon | 0.62 | Not Significant | | Zambia | 4.15 | Not significant | |
| Namibia | 0.97 | Not Significant | | Norway | 0.04 | Not significant | |
| Senegal | 0.62 | Not Significant | | Australia | 0.38 | Not significant | |
| Turkmenistan | 0.58 | Not Significant | | Sweden | 0.04 | Not significant | |
| Timor-Leste | 5.13 | Not Significant | |  |  |  |  |
| Kazakhstan | 0.58 | Not Significant | |  |  |  |  |
| Zambia | 1.38 | Not Significant | |  |  |  |  |
| ***Increasing trend*** | | | | | | | |
| Guam | 7.83 | 0.004 | <0.05 | Dominica | 11.21 | 0.001 | <0.05 |
| Malawi | 1.38 | 0.004 | <0.01 | Peru | 11.19 | 0.002 | <0.001 |
| South Sudan | 1.38 | 0.005 | <0.001 | South Sudan | 4.15 | 0.003 | <0.01 |
| Somalia | 1.38 | 0.005 | <0.001 | Guam | 23.60 | 0.005 | <0.001 |
| Bangladesh | 2.97 | 0.006 | <0.001 | Malawi | 4.16 | 0.005 | <0.001 |
| Uganda | 1.38 | 0.006 | <0.001 | Suriname | 11.21 | 0.005 | <0.05 |
| Madagascar | 1.38 | 0.007 | <0.01 | Somalia | 4.16 | 0.005 | <0.001 |
| Palau | 7.84 | 0.007 | <0.001 | Bangladesh | 8.97 | 0.005 | <0.001 |
| Belize | 3.73 | 0.008 | <0.05 | Uganda | 4.16 | 0.005 | <0.001 |
| Jamaica | 3.72 | 0.008 | <0.001 | Madagascar | 4.16 | 0.007 | <0.01 |
| United States Virgin Islands | 3.71 | 0.009 | <0.01 | Palau | 23.64 | 0.007 | <0.001 |
| Chad | 0.62 | 0.010 | <0.01 | Belize | 11.24 | 0.008 | <0.05 |
| Democratic Republic of the Congo | 0.98 | 0.010 | <0.05 | United States Virgin Islands | 11.19 | 0.009 | <0.01 |
| Canada | 0.15 | 0.015 | <0.01 | Chad | 1.88 | 0.009 | <0.001 |
| Greenland | 0.15 | 0.018 | <0.01 | Jamaica | 11.21 | 0.009 | <0.001 |
| Democratic People's Republic of Korea | 4.53 | 0.018 | <0.001 | Democratic Republic of the Congo | 2.96 | 0.010 | <0.05 |
| Republic of Korea | 0.08 | 0.034 | <0.01 | Canada | 0.45 | 0.010 | <0.01 |
| Argentina | 0.08 | 0.035 | <0.001 | Democratic People's Republic of Korea | 13.75 | 0.017 | <0.001 |
| Brunei Darussalam | 0.08 | 0.037 | <0.05 | Greenland | 0.45 | 0.018 | <0.001 |
| Mali | 0.67 | 0.042 | <0.001 | Republic of Korea | 0.23 | 0.034 | <0.01 |
| Japan | 0.08 | 0.043 | <0.05 | Brunei Darussalam | 0.23 | 0.036 | <0.05 |
| Uruguay | 0.08 | 0.044 | <0.001 | Argentina | 0.23 | 0.037 | <0.001 |
| Monaco | 0.01 | 0.057 | <0.01 | Mali | 2.02 | 0.037 | <0.001 |
| Germany | 0.01 | 0.057 | <0.05 | Japan | 0.24 | 0.043 | <0.05 |
| Switzerland | 0.01 | 0.058 | <0.001 | Uruguay | 0.23 | 0.044 | <0.001 |
| Greece | 0.01 | 0.059 | <0.001 | Germany | 0.02 | 0.052 | <0.05 |
| Denmark | 0.01 | 0.061 | <0.001 | Monaco | 0.03 | 0.058 | <0.01 |
| Malta | 0.01 | 0.061 | <0.001 | Switzerland | 0.03 | 0.059 | <0.001 |
| Netherlands | 0.01 | 0.061 | <0.001 | Greece | 0.03 | 0.060 | <0.001 |
| Ireland | 0.01 | 0.061 | <0.001 | Denmark | 0.03 | 0.061 | <0.001 |
| Luxembourg | 0.01 | 0.062 | <0.001 | Malta | 0.03 | 0.062 | <0.001 |
| Finland | 0.01 | 0.062 | <0.001 | Netherlands | 0.03 | 0.062 | <0.001 |
| Israel | 0.01 | 0.062 | <0.001 | Ireland | 0.03 | 0.062 | <0.001 |
| Iceland | 0.01 | 0.062 | <0.001 | Luxembourg | 0.03 | 0.062 | <0.001 |
| Spain | 0.01 | 0.063 | <0.001 | Finland | 0.03 | 0.062 | <0.001 |
| France | 0.01 | 0.065 | <0.001 | Israel | 0.03 | 0.063 | <0.001 |
| Austria | 0.01 | 0.065 | <0.001 | Iceland | 0.03 | 0.063 | <0.001 |
| Norway | 0.01 | 0.065 | <0.001 | Spain | 0.03 | 0.064 | <0.001 |
| San Marino | 0.01 | 0.066 | <0.001 | Cyprus | 0.03 | 0.065 | <0.001 |
| Australia | 0.13 | 0.066 | <0.01 | Austria | 0.03 | 0.065 | <0.001 |
| Sweden | 0.01 | 0.066 | <0.001 | France | 0.03 | 0.066 | <0.001 |
| Ethiopia | 2.53 | 0.105 | <0.001 | San Marino | 0.03 | 0.066 | <0.001 |
| Italy | 0.04 | 0.122 | <0.05 | Ethiopia | 7.53 | 0.110 | <0.001 |
| Romania | 1.44 | 0.146 | <0.001 | Italy | 0.12 | 0.119 | <0.05 |
| U.K. of Great Britain and Northern Ireland | 0.01 | 0.262 | <0.001 | Romania | 4.33 | 0.138 | <0.001 |
| Mexico | 3.40 | 0.560 | <0.001 | U.K. of Great Britain and Northern Ireland | 0.04 | 0.265 | <0.001 |
|  |  |  |  | Mexico | 10.26 | 0.555 | <0.001 |

**Supplementary Figure 5: Spatial Clustering and Moran’s *I^[1]^***

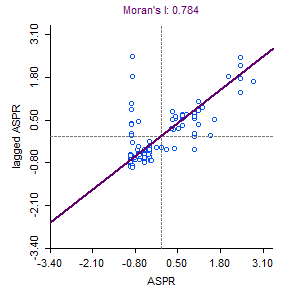

**Supplementary Table 2: Association of scabies rates with socioeconomic factors**

|  | ln(Age-standardized incidence rate) | | | ln(Age-standardized prevalence rate) | | |
| --- | --- | --- | --- | --- | --- | --- |
| Covariates | *β* | *p*-value | 95% CI | *β* | *p*-value | 95% CI |
| (Intercept) | 3.355 | 0.012 | (0.02 to 0.97) | 2.194 | 0.098 | (-0.40 to 4.79) |
| ^$^Latitude* | -0.057 | <0.001 | (0.74 to 5.96) | - 0.057 | <0.001 | (-0.07 to -0.04) |
| % urban population* | -0.028 | <0.001 | (-0.07 to -0.04) | -0.028 | <0.001 | (-0.04 to -0.01) |
| Life expectancy at birth | -0.007 | 0.813862 | (-0.04 to -0.01) | -0.006 | 0.834 | (-0.06 to 0.04) |
| ln(% of unemployment) | 0.107 | 0.43926 | (-0.06 to 0.04) | 0.108 | 0.433 | (-0.16 to 0.38) |
| ln(population density) | -0.106 | 0.224919 | (-0.16 to 0.37) | -0.106 | 0.223 | (-0.27 to 0.06) |
| ln(GNI) | 0.129 | 0.530124 | (-0.27 to 0.06) | 0.129 | 0.527 | (-0.27 to 0.53) |

^$^absolute value. **p<*0.001, CI: confidence interval.

**Supplementary Figure 6: Model diagnostics**

| (a) **Fitting of model residuals** | (b) **Histogram of model residuals** |
| --- | --- |
| 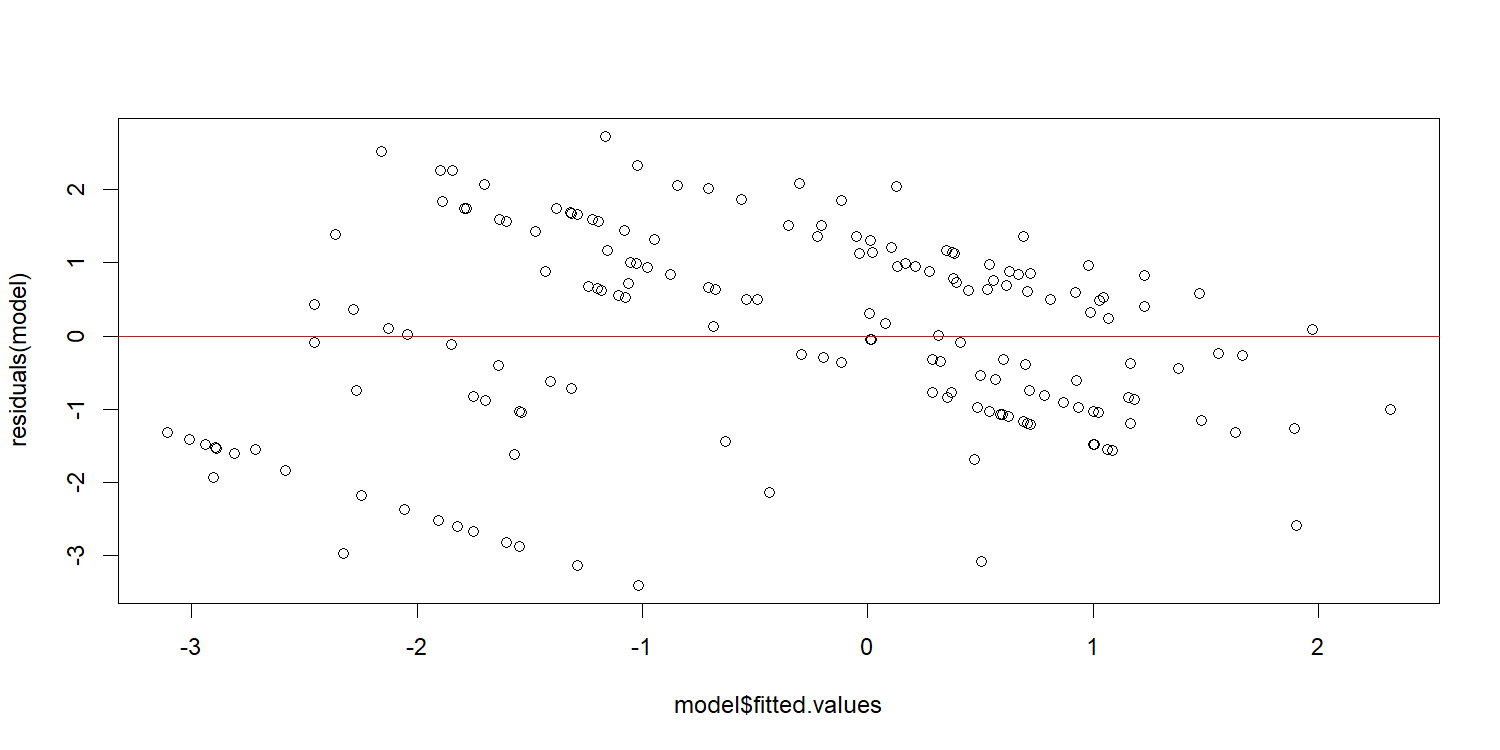 | 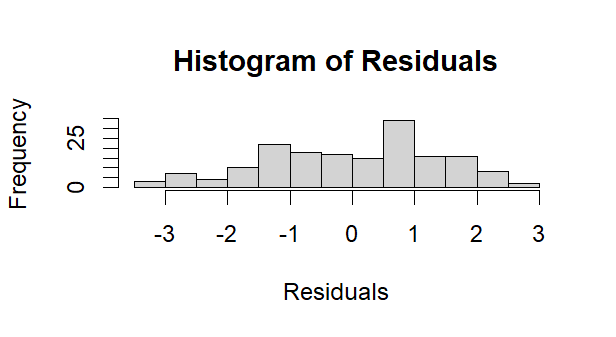 |
| **(c) QQ Plot of the model** | |
| 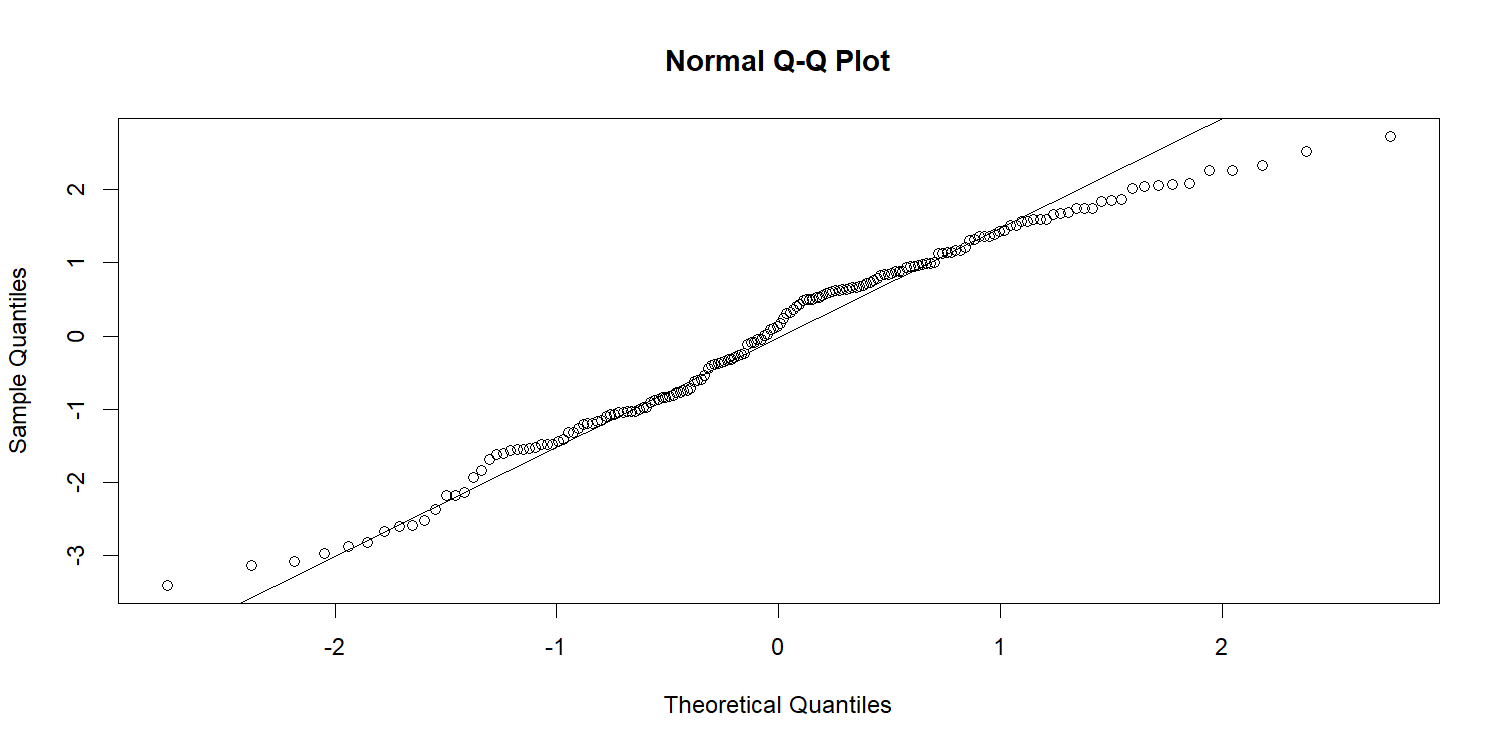 | |

1. A Moran’s *I* of 0.784 indicates strong clustering and positive autocorrelation
